# Host and microbiome features of secondary infections in lethal covid-19

**DOI:** 10.1101/2022.02.18.22270995

**Authors:** Martin Zacharias, Karl Kashofer, Philipp Wurm, Peter Regitnig, Moritz Schütte, Margit Neger, Sandra Ehmann, Leigh M. Marsh, Grazyna Kwapiszewska, Martina Loibner, Anna Birnhuber, Eva Leitner, Andrea Thüringer, Elke Winter, Stefan Sauer, Marion J. Pollheimer, Fotini R. Vagena, Carolin Lackner, Barbara Jelusic, Lesley Ogilvie, Marija Durdevic, Bernd Timmermann, Hans Lehrach, Kurt Zatloukal, Gregor Gorkiewicz

## Abstract

Secondary infections contribute significantly to covid-19 mortality but host and microbial factors driving this sequel remain poorly understood. We performed an autopsy study of 20 covid-19 cases and 14 controls from the first pandemic wave. Autopsies combined with microbial cultivation and deep RNA sequencing (RNAseq) allowed us to define major organ pathologies and specify secondary infections. Lethal covid-19 segregated into two main death causes separating cases with either dominant diffuse alveolar damage (DAD) or secondary infections of lungs. Lung microbiome changes were profound in covid-19 showing a reduced biodiversity and increased presence of prototypical bacterial and fungal pathogens in cases with secondary infections. Deep RNAseq of lung tissues distinctly mirrored death causes and cellular deconvolution stratified DAD cases into subgroups with different cellular compositions. Myeloid cells, including macrophages, and complement C1q activation were found to be strong stratifying factors suggesting a pathophysiological link possibly leading to tolerance in DAD subgroups. Moreover, several signs of immune-impairment were evident in covid-19 lungs including strong induction of inhibitory immune-checkpoints. Thus, our study highlights profound alterations of the local immunity in covid-19, wherein immune-impairment leads to reduced antimicrobial defense favoring the development of secondary infections on top of SARS-CoV-2 infection.

## Introduction

Covid-19 originates from infection of the upper respiratory tract with SARS-CoV-2, which can progress into severe acute lung injury (ALI). Based on the tissue-typic expression of the viral host-entry receptor ACE2 and certain proteases (e.g. TMPRSS2) facilitating cellular uptake, also other organs like the kidney could be directly infected [1]. In addition, severe disturbance of immune and coagulation systems during covid-19 lead to a multifaceted disease with variable multi-organ damages [2]. A consistent finding in severe covid-19 is initial immune hyperactivation (called “cytokine storm”) leading to subsequent immune exhaustion, a phenomenon also known in other severe infections [3-6]. Consequently, secondary infections which develop on top of SARS-CoV-2 infection contribute significantly to covid-19 mortality similar to severe influenza [7]. Curiously, the pathophysiology leading to the development of secondary lung infections is generally poorly understood. We performed an autopsy study of 20 consecutive covid-19 patients, who died during the first pandemic wave. Full autopsies were performed and various specimen types were collected for tissue-based investigations, molecular measures including deep sequencing and cultivation of virus and other microbes. Integrating all the information gained from this “holistic” autopsy approach allowed us to gain a deeper understanding of host and microbial factors contributing to secondary infections as a major sequel of lethal covid-19.

### Autopsy cohort, SARS-CoV-2 body distribution and genotyping

Twenty consecutive covid-19 patients were examined post-mortem (Figure S1). Thirteen cases were males and 7 were females, their age ranged from 53 to 93 years (median: 79 years). All had multiple comorbidities typically prevalent in severe covid-19. In addition, 14 age-matched non-covid-19 controls who died within the same time period were included for comparisons (Tables S1 and S2). Patients were tested for SARS-CoV-2 tissue distributions by quantitative RT-PCR (target: nucleocapsid-gene) and most positive samples with the highest viral loads originated from the respiratory tract, followed by myocardium, liver, kidney and pleural effusions. Other tissues and body liquids were positive only in single cases or tested overall negative (Figure 1A). Notably, deep RNAseq generated from lung tissues revealed SARS-CoV-2 transcripts in each covid-19 case, including the four qRT-PCR negative ones, showing increased sensitivity of deep transcriptomic analysis (127±29 million reads were generated per sample on average; Figure 1B). The viral genome was entirely captured by deep RNAseq from lung tissues yielding more plus-strand reads (mean: 37.89 reads per million; range: 0.02-131,165.41) than minus-strand reads (mean: 1.81 reads per million; range: 0-484.81; Figure 1C). In addition, 11 SARS-CoV-2 strains could be cultivated from post-mortem lung tissues using Vero cells (Table S3). Successful virus cultivation significantly correlated with abundance of SARS-CoV-2 reads (Figure 1D and S2).

**Figure 1.**
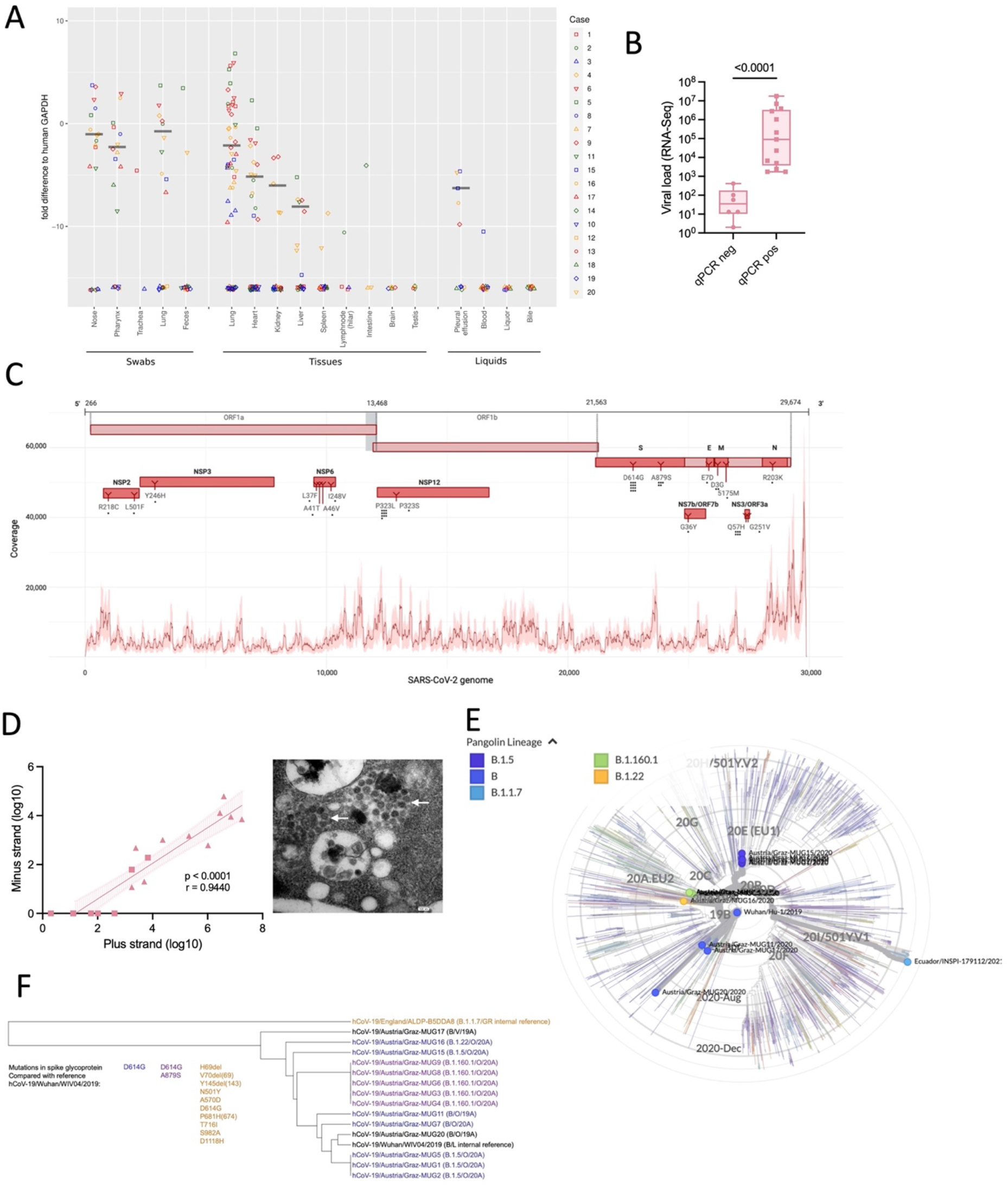
SARS-CoV-2 tissue distributions, genotyping and virus cultivation. (A) SARS-CoV-2 loads (compared to human glyceraldehyde 3-phosphate dehydrogenase, GAPDH) and tissue distributions derived from post mortem sampling (median highlighted). Case numbers are given on the right. (B) Significant association of qRT-PCR positivity (n-gene) with viral loads determined by RNAseq of lung tissues (Mann-Whitney test). (C) Distribution of viral reads generated from lung tissues along the SARS-CoV-2 genome. Cumulative coverage of plus and minus strand transcripts is shown (median in bold). Identified nucleotide and amino-acid changes in comparison to the Wuhan reference strain are indicated. (D) Correlation of SARS-CoV-2 plus and minus strand reads with cultivation (Spearman correlation). Triangles specify cultivation-positive samples. EM picture showing viral particles in Vero CCL-81 cells (arrows). (E)Cladogram showing detected virus genotypes within a global context. The Wuhan reference strain (center) and the UK variant B.1.1.7 (Ecuador/INSPI-179112/2021) are included for comparisons. The pangolin lineage designation is used to specify viral genotypes. (F)Dendrogram showing detected viral genotypes. Corresponding mutations in the S protein are indicated and virus strains are color coded accordingly.

SARS-CoV-2 genotyping facilitated by PCR and sequencing directly from autopsy specimens yielded 14 complete viral genomes (Table S4). Nine different sequence variants were detected showing up to 12 nucleotide changes compared to the reference (SARS-CoV-2 Wuhan-Hu-1; total genome size 29,903 bp; Table S5). Strains corresponded to the pangolin lineages B.1.22, B1.5, B and B.1.160.1, respectively (clades 19A and 20A), representing the dominant genotypes of the first pandemic wave (Figure 1E). Twelve strains harbored a D614G mutation in the spike (S) protein, which leads to increased viral transmissibility and, therefore, this genotype superseded the wild-type strain already early in the pandemic [8]. We identified also 2 viral clusters in our cohort, cluster 1 (case 3, 4, 6, 8, and 9) and cluster 2 (case 1, 2, and 5), respectively (Figure 1F).

Notably, cases 6, 8 and 9 from cluster 1 originated from the same residential care home and all cases from cluster 2 stayed in the same hospital ward prior to covid-19. Thus, it is very likely that these individuals were infected from the same sources and/or transmission occurred.

### Major organ pathologies and death causes

Lungs showed the dominant pathologies in relation to covid-19, only one case (#1) presented with acute myocardial infarction as the ascribed death cause. Diffuse alveolar damage (DAD), the histopathological representation of ALI, in a patchy distribution and often prevalent in multiple lung segments was the major finding in 11 cases. Early exudative stages and later organizing stages of DAD were found within the same patient together, often adjacent to nearly normal or less affected parenchyma indicating ongoing tissue damage (Figures 2A and S3-S5). Also, a significant positive correlation of SARS-CoV-2 loads from nasopharyngeal tissues compared to lungs was found (Figure 2B), which suggests active seeding of infectious particles from the upper respiratory tract likely via micro-aspiration into the lungs [1]. Microscopic features of lungs were extensively assessed (see methods for details of scoring histopathological changes) to specify and grade the severity of lesions and also to capture the heterogeneity of different lung pathologies. Features greatly varied between cases and no pattern clearly correlated with disease duration (defined as the interval between the first SARS-CoV-2 positive PCR and death) or viral loads (Figure 2C). Importantly, the clearest discriminating feature of cases was the presence of neutrophilic granulocytes, indicative of secondary infections (i.e. “pneumonia”), in comparison to DAD.

**Figure 2.**
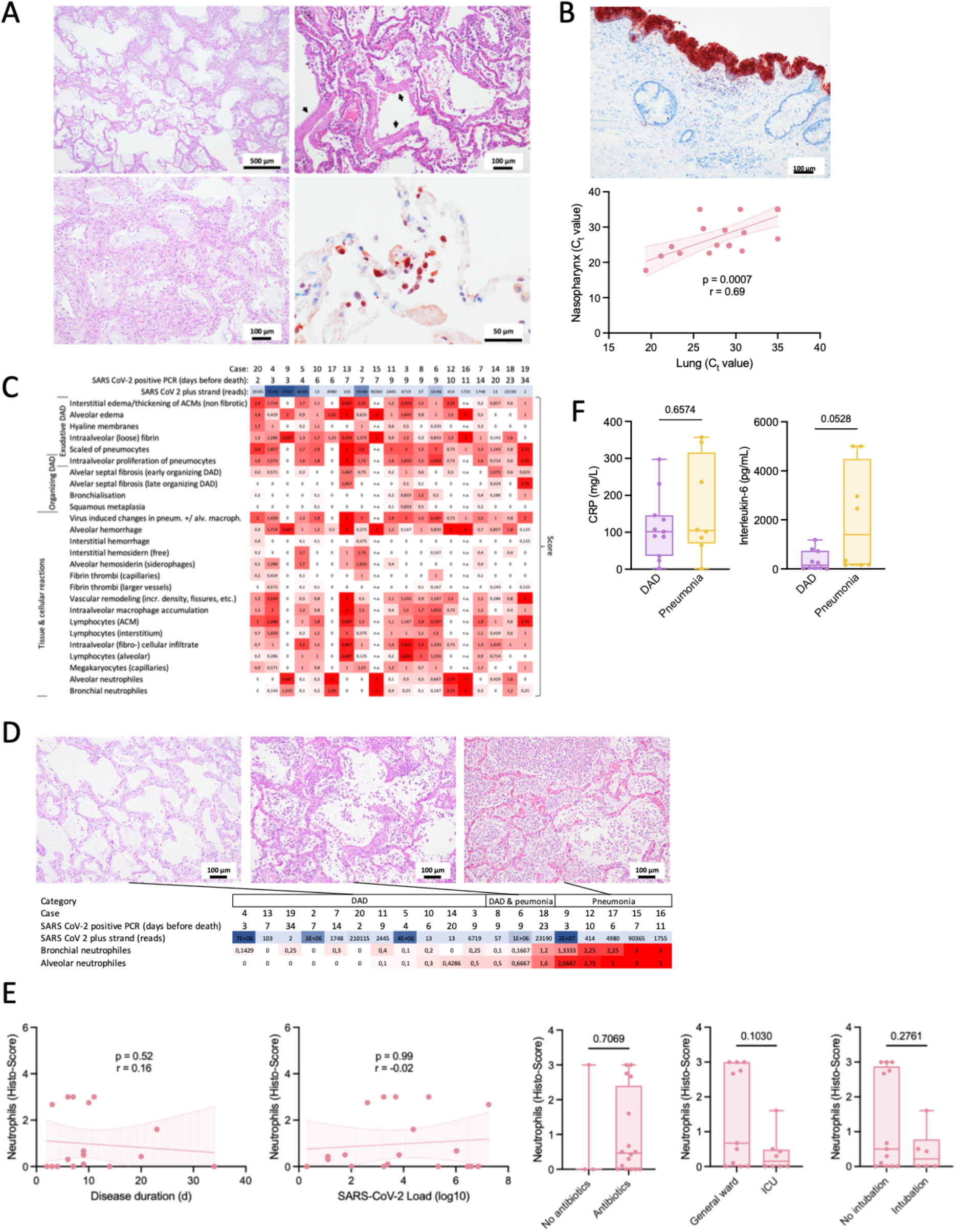
Lung pathology of lethal covid-19 stratifies into diffuse alveolar damage and pneumonia. (A) Histological representation of diffuse alveolar damage (DAD) in lungs. A patchy representation of DAD is shown (left). Hyaline membranes (arrows) as a hallmark lesion of early DAD (top right). Immunohistochemical detection (nucleoprotein antibody) of SARS-CoV-2 infected pneumocytes (bottom right). (B) (Top) Immunohistochemical detection of SARS-CoV-2 infected respiratory epithelium of the nasopharynx. (Bottom) Correlation between SARS-CoV-2 loads in the nasopharyngeal mucosa and lung tissue determined by qRT-PCR (Spearman correlation). (C) Scoring of prevalent histopathology patterns in lungs. Cases are ordered according to duration of disease. (D) Main discrimination of lung pathology according to DAD and pneumonia patterns. Cases are ordered according to alveolar neutrophil counts. (E) Correlation analyses of neutrophil abundance and clinical parameters (Spearman correlation; Mann-Whitney test). (F) Serum C-reactive protein (CRP) and interleukin-6 (IL-6) levels (Mann-Whitney test) in DAD versus pneumonia cases.

Three cases showed DAD superimposed with acute inflammation and 5 cases showed mainly pneumonia as the dominant pathology, wherein DAD was only focally visible overlaid with dense inflammation. Altogether, 16 cases showed neutrophilic granulocytes present in bronchi, bronchioli or alveoli suggestive of secondary infections. Thus, lung histopathology in lethal covid-19 could be stratified into DAD, DAD superimposed with pneumonia and dominating pneumonia (Figures 2D and S3). Noteworthy, neither disease duration nor viral loads correlated with presence of neutrophils, nor did any other clinical parameter (Figures 2E and S6), however, pneumonia cases showed increased IL-6 levels compared to pure DAD cases, whereas CRP levels did not differ between categories (Figure 2F). Other organs showed features of preexisting comorbidities including arteriosclerosis, hypertension and diabetes, especially in the kidneys, wherein SARS-CoV-2 could be detected in tubular epithelia by positive immunohistochemistry (Figure S7). Heart and liver specimens revealed no clear evidence of direct SARS-CoV-2 carriage nor features of myocarditis or hepatitis (Figures S8 and S9). A detailed summary of organ histopathologies is given in the supplementary appendix (Table S6).

### Lung microbiome alterations and secondary-infections in lethal covid-19

The lung microbiome is altered in DAD and thought to be a relevant factor for the development of secondary infections [9, 10]. RNAseq data from lung tissues was screened for microbial sequences and bacterial (16S rRNA gene) and fungal (internal transcribed spacer, ITS) marker genes were amplified to additionally specify microbial changes. On average 6573.33±2552.32 (MW±SD) reads per million (rpm) per sample were not human in RNAseq and likely of microbial origin, of those 2.02±4.00% and 0.03±0.05% could be clearly annotated to specific microbes with different microbial annotation pipelines (Figure 3A). Excluding SARS-CoV-2 reads, which were the dominant microbial component in several cases (range: 0.01 – 131218.36 rpm), bacterial sequences were dominant, significantly increased in covid-19 cases with pneumonia compared to DAD and controls. Fungal and viral sequences other than SARS-CoV-2 were also significantly enriched in covid-19 cases with pneumonia (Figure 3B). Number of bacterial reads significantly correlated with neutrophil scores suggesting that their presence is a sign of secondary infections, however, the post-mortem interval did not, precluding a strong influence of post-mortal bacterial overgrowth in our investigation (Figure 3C). Bacteria are assumed to be the dominant microbiome component in lungs [11]. Analysis based on the bacterial 16S rRNA gene marker showed that richness was significantly decreased in the DAD and pneumonia cases of covid-19 compared to controls indicating an overall reduced biodiversity (Figure 3D). In contrast, evenness was significantly decreased in the pneumonia group of covid-19 only, suggesting a dominance of certain taxa, possibly representing the agents of secondary infections (pairwise Kruskal-Wallis; *p<0.05,**p<0.005). Principal component analysis (PCA) clearly separated controls from covid-19 cases with DAD and cases with pneumonia indicating significantly different bacterial community compositions (Figure 3D). Lung tissues were also cultured for bacteria and fungi and both, covid-19 cases and controls, yielded cultivable microorganisms but in different quantities and taxonomic constellations (Table S7).

**Figure 3.**
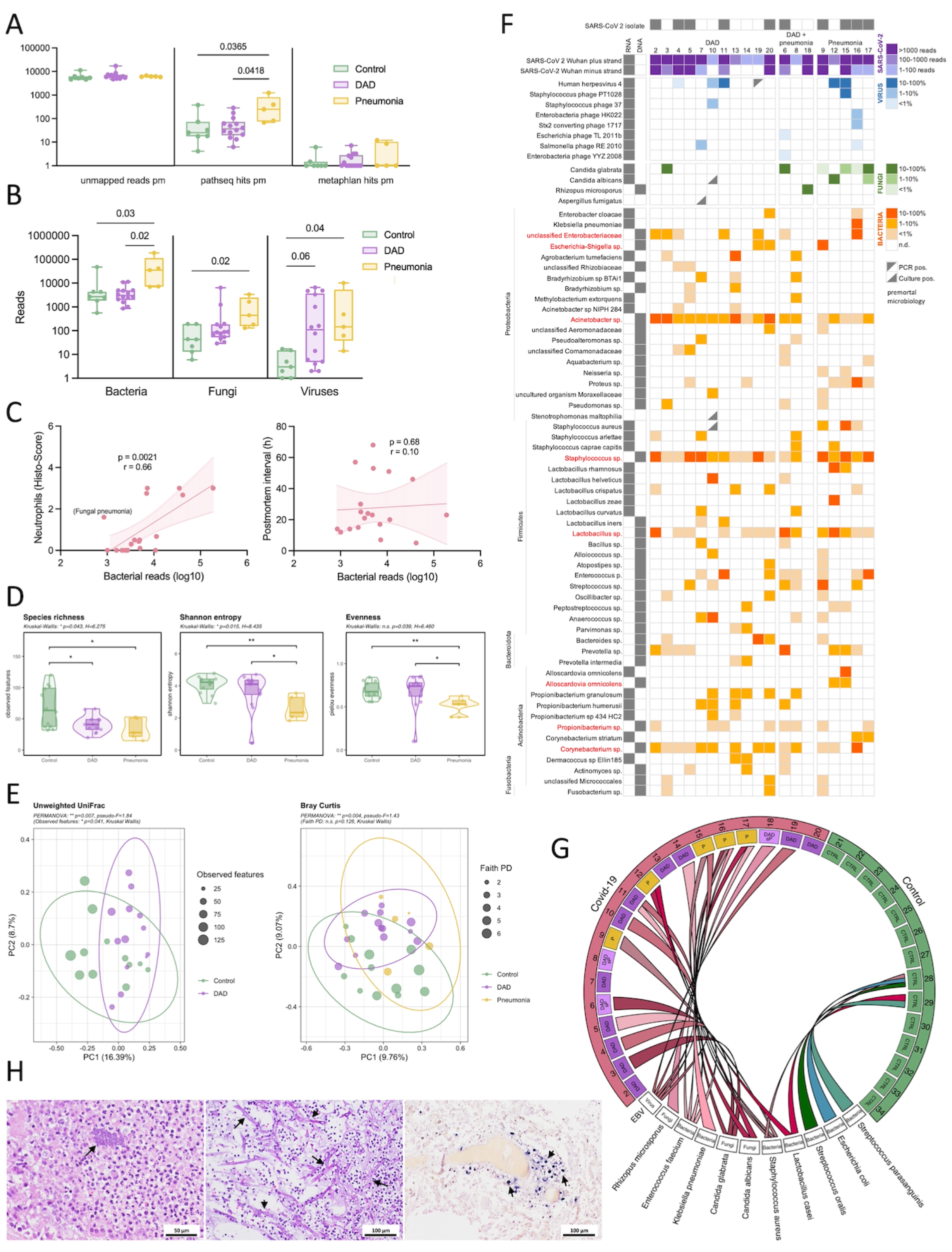
Microbiome alterations and agents of secondary infections in covid-19 lungs. (A) Annotation of non-human transcripts to microbial sequences with PathSeq and MetaPhlAn, respectively. (B) Significantly increased bacterial, fungal and viral reads in the pneumonia category of covid-19 (PathSeq annotation, Kruskal-Wallis test). (C) Bacterial reads significantly correlate with neutrophil counts but not with the post-mortem interval (Spearman correlation). (D) Richness and evenness in the bacterial component of the lung microbiome (based on the 16S rRNA gene marker). (E) Beta-diversity analysis (PCA based on unweighted UniFrac and Bray Curtis distance) clearly separates DAD and pneumonia cases of covid-19 from controls (16S rRNA gene). (F) Summary of bacterial, fungal and viral microbes prevalent in covid-19 lungs. Shown are microbes detected by cultivation, RNA and/or DNA sequencing.(G)Dominant pathogens causing secondary infections in covid-19 lungs compared to controls (summary of cultivation and deep sequencing). (H) Microscopic representation (H&E) of bacterial (left, case #16) and fungal (middle, case #18) pathogens in lung tissues. Epstein Barr virus RNA positivity in lung tissue (EBV RNA in-situ hybridization, case #11).

Finally, we integrated RNAseq, 16S, ITS and culture data to define dominant pathogens, most likely representing the agents of secondary infections and to account for the different samples used for microbial identifications in the light of the patchy disease representations likely impacting the microbial repertoires (Figure S10, Table S8). Dominant pathogens were defined if they were dominant in the RNA and/or DNA data (representing > 10% of microbial reads excluding SARS-CoV-2) and if they also yielded a reasonable culture growth (≤10^4^ cfu/ml). Dominant taxa were typical agents of pulmonary secondary infections like *Staphylococcus aureus, Enterococcus faecium* or *Klebsiella pneumoniae*, as well as fungi like *Candida* spp. or the mold *Rhizopus microsporus* identified in one case (#18). Often multiple pathogens were found simultaneously indicating polymicrobial infections (e.g. in case #16 wherein *K. pneumonia, S. aureus* and *C. glabrata* were cultivated in reasonable amounts and were also captured by RNAseq). In addition, 5 covid-19 cases yielded transcripts of Epstein Barr virus (EBV), which were also detectable by RNA *in-situ* hybridization of lung tissues but not in controls (Figure 3H). EBV often emerges due to endogenous reactivation in the context of impaired immunity [12]. Control cases yielded microbial sequences and cultivable microbes in lower quantity and they often belonged to known contaminants like *Lactobacillus* sp. or *Propionibacterium* sp. (Table S8). In summary, the lung microbiome in covid-19 shows a reduced taxonomic richness but harbors a diverse spectrum of bacterial and fungal pathogens typically associated with secondary lung infections. Prominent pathogens like *S. aureus, Klebsiella* or *Candida* spp. are also known agents of secondary infection in influenza, SARS and MERS [13, 14]. Notably, secondary infections were rarely detected ante-mortem in our cohort (Table S8). The presence of poly-microbial infections and the relatively high proportion of EBV positivity suggests an overall impaired immunity in covid-19 lungs.

### The lung metatranscriptome mirrors the major death categories DAD and pneumonia

Deep RNAseq of lung tissue revealed 4,547 differentially expressed genes between covid-19 cases and controls (adj. P<0.05). Hierarchical clustering indicated depleted (cluster 1) or enriched (cluster 2) genes in covid-19 compared to controls (Figure 4A). Pathway analysis indicated impaired central cellular functions within mRNA metabolism, post-translational protein modification, the respiratory chain, VEGFA signaling and extracellular matrix organization in covid-19. Enriched pathways consisted mainly of innate and adaptive immune functions, neutrophil degranulation, cytokine signaling as well as complement activation (Figure 4B). Overall, these data confirm profound and complex transcriptional alterations in covid-19 lung tissue [15-17]. Unsupervised principal components analysis (PCA) of differentially expressed genes clearly separated covid-19 samples on principal component 1 (PC1) from controls but also clearly separated pneumonia samples from pure DAD cases (Figure 4C). Comparison of differentially expressed genes between these major death categories indicated that the major discriminator from controls was DAD showing 3862 unique differentially expressed genes (adj. P < 0.05 and abs LFC >= 0.58) followed by pneumonia with 1673 unique differentially expressed genes (Figure 4D). DAD and pneumonia differed by only 226 differentially expressed genes. Notably, amongst the top 50 differential expressed genes enriched in pneumonia cases several macrophage markers were evident, including the receptor *ADGRE1* (murine homolog F4/80) as top-hit, the interleukin-1 receptor-associated kinase-like 2 (*IRAK2*) or *PSTPIP2*, which is involved in macrophage polarization (Figure 4E).

**Figure 4.**
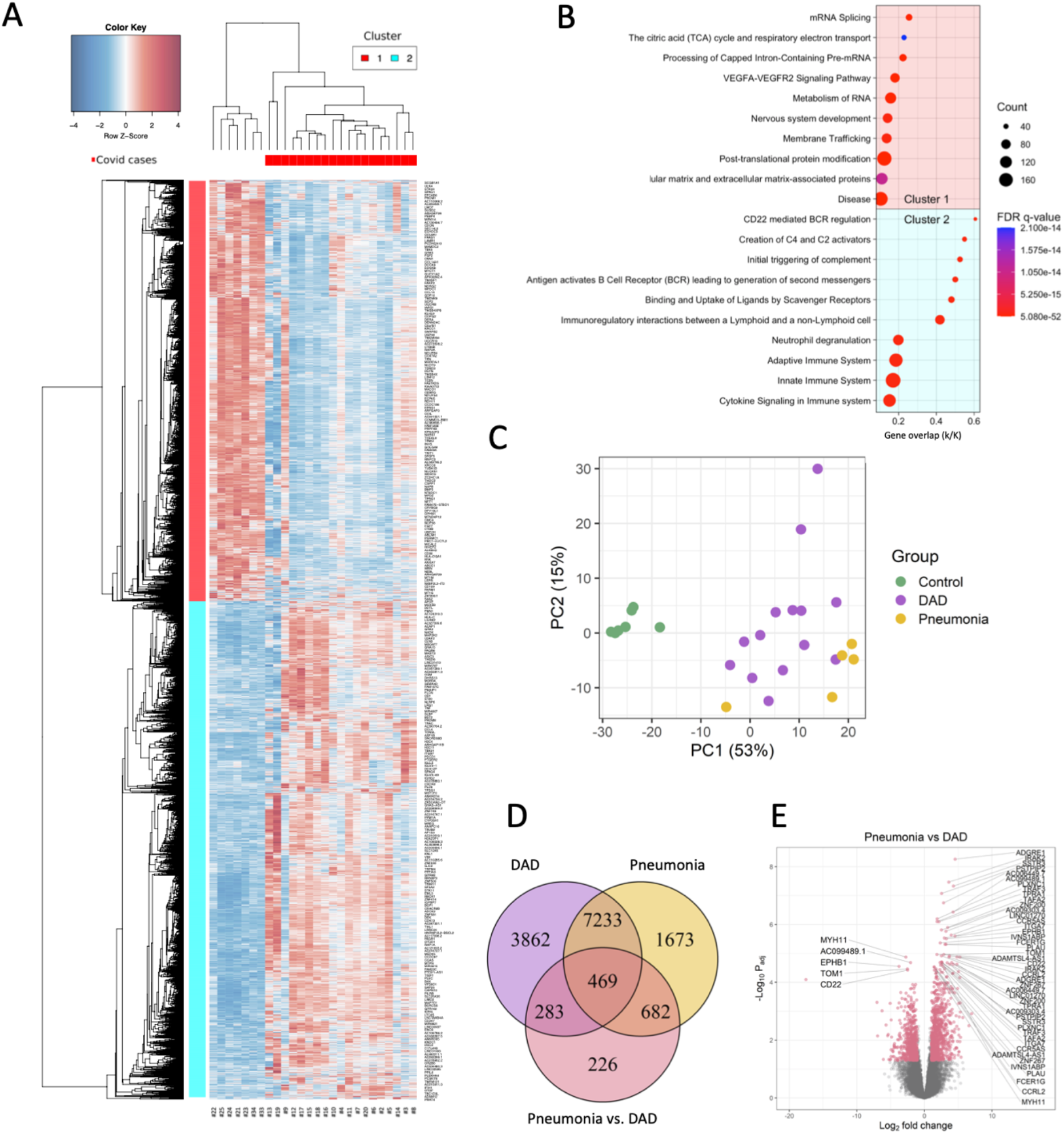
The lung metatranscriptome mirrors the major death categories DAD and pneumonia. (A) Hierarchical clustering shows depleted (cluster 1) and enriched (cluster 2) genes (n=4,547; adj. P<0.05) in lung tissue of covid-19 cases compared to controls. (B) Gen set enrichment analysis (canonical pathways) of major depleted (top) and enriched (bottom) pathways in covid-19 lungs. (C) PCA based on differentially expressed genes clearly discriminates DAD cases and cases with secondary pneumonia of covid-19 from controls. (D) Venn diagram specifying differentially expressed genes in DAD as the major discriminator followed by pneumonia (adj. P<0.05, LFC≥0.58). (E) Volcano plot showing the top 25 significantly deregulated genes in secondary pneumonia versus DAD. Several macrophage genes are increased.

Thus, macrophages seem to be implicated in covid-19 secondary infections. In summary, deep transcriptomic analyses specified multiple dysregulated processes in covid-19, including vascular and coagulation systems, connective tissue remodeling as well as activated immunity and complement [18]. Similar to histopathology, the major discriminator from controls based on gene expression was DAD followed by pneumonia likely mirroring the development of secondary infections on top of ALI caused by the virus.

### Cellular deconvolution subgroups covid-19 lung pathology

Cellular compositions were inferred from deep RNAseq data by using xCell [19]. Hierarchical clustering based on cellular compositions clearly separated samples into four distinct groups. Group 1 (“control”) consisted only of control cases and was related to group 2 (“DAD1”) consisting of covid-19 cases with pure DAD (in addition to one control case #22). Group 3 (“DAD2”) contained also DAD cases, including one sample with the histological category DAD and secondary pneumonia. This group was related to group 4 (“pneumonia”) composed of all pneumonia cases, in addition to 3 DAD cases and the two remaining DAD cases with secondary pneumonia (Figure 5A). Noteworthy, neither disease duration nor SARS-CoV-2 loads clearly correlated with a specific grouping. Cell types discriminating these groups showed a specific assembly (Figure 5B). Cluster 1 consisted mainly of vascular and stromal cell types like endothelial cells, pericytes and fibroblasts, enriched in “DAD1” and “DAD2". Top enriched genes in this cluster were certain collagen genes, the vascular transcription factor *sox 18*, the basement membrane protein *ladinin-1* or the endothelial protein *stabilin-1* (Figure 5C). Cell types of cluster 2 consisted mainly of structural and stromal cells, in addition to certain immune and blood cell types and they were overall reduced in covid-19.

**Figure 5.**
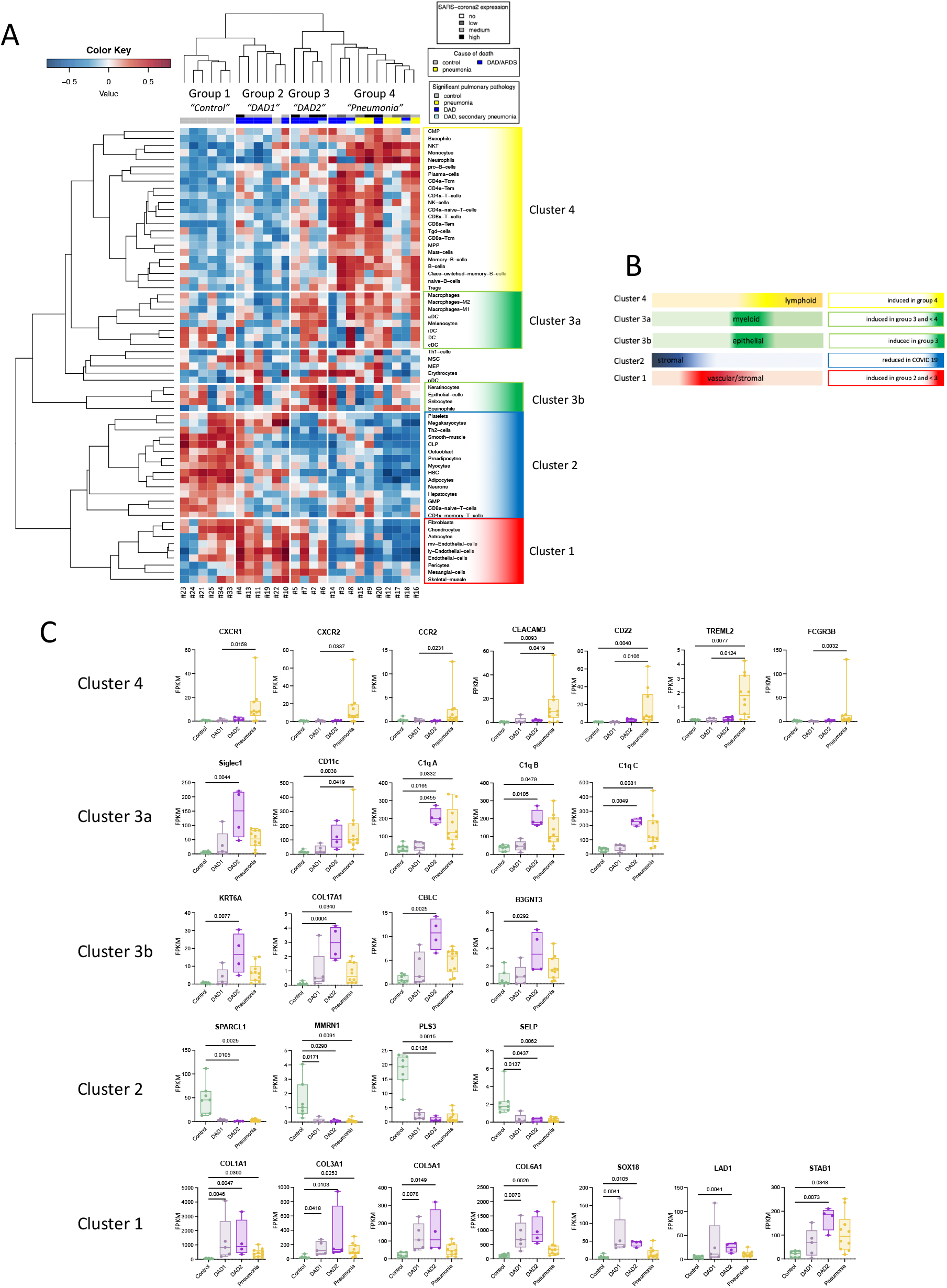
Cellular deconvolution stratifies lung pathology sub-groups. (A) Hierarchical clustering based on cell-type enrichments derived from xCell analysis indicates a specific grouping of samples. (B) Scheme indicating cell clusters which discriminate different groups.(C) Top induced genes in the respective cell clusters determining the specific grouping (Kruskal-Wallis test).

Top down-regulated genes included the extracellular matrix proteins *sparc-like 1, multimerin-1* and *plastin-3* or the cell adhesion molecule *p-selectin* important for the recruitment of leukocytes typically prevalent on activated endothelial cells and platelets (Figure 5C). Together these alterations highlight the vascular and connective tissue changes emerging during DAD development (Figure 2B) [20, 21]. Cell types of cluster 3, which were dominantly induced in “DAD2” and to a lesser extent in “pneumonia” showed enrichment of myeloid (cluster 3a) and epithelial cell types (cluster 3b). Top induced genes in cluster 3a were the myeloid cell specific genes *siglec-1, CD11c* and complement factor *C1q*. Top induced genes in cluster 3b consisted of *keratin 6A, collagen XVII*, the tyrosine kinase signaling protein *cbl-c* and beta-1,3-N-acetylglucosaminyltransferase 3 (*b3gnt3*), typically expressed in epithelia and also involved in lymphocyte trafficking and homing. Cell types in cluster 4, strongly increased in “pneumonia” consisted of different leukocyte classes including B-, T-cells and (neutrophilic) granulocytes. Top induced genes consisted of the interleukin 8 receptor genes *cxcr1* and *cxcr2*, the chemokine receptor type 2 (*CCR2*), *CEACAM3, CD22* (B cell marker), and the cell surface receptors *TREML2* and *FCGR3B*.

In summary, cellular deconvolution clearly sub-stratified the major categories DAD and pneumonia of covid-19 lung pathology. Noteworthy, DAD subclustered into two different groups, one showing mainly induction of vascular and stromal cell elements (“DAD1”), the other dominant induction of genes related to myeloid and epithelial cells (“DAD2”), and this subgroup showed more commonalities with the pneumonia group.

### Macrophages, complement c1q and immune impairment in covid-19 lungs

Myeloid cells including macrophages play a central role in the pathogenesis of DAD [22-24], and bronchioloalveolar lavage fluids (BALF) of patients with severe covid-19 show high proportions of macrophages [15, 25]. We confirmed significantly increased macrophages in covid-19 lungs by CD163 Immunohistochemistry, a M2-type macrophage marker (Figure 6A), corroborating a recent proteomic study wherein CD163 was found amongst the most induced proteins in lungs and spleens derived from covid-19 autopsies [18]. Deconvolution indicated both M1- and M2-type macrophages significantly enriched predominantly in “DAD2” whereas monocytes were mainly induced in the “pneumonia” group (Figure 6B). Increased CD163 positive macrophages gathering around virus positive cells were recently shown also in a macaque model of SARS-CoV infection, indicating that infected pneumocytes may lead to macrophage recruitment in coronavirus infections [26].

**Figure 6.**
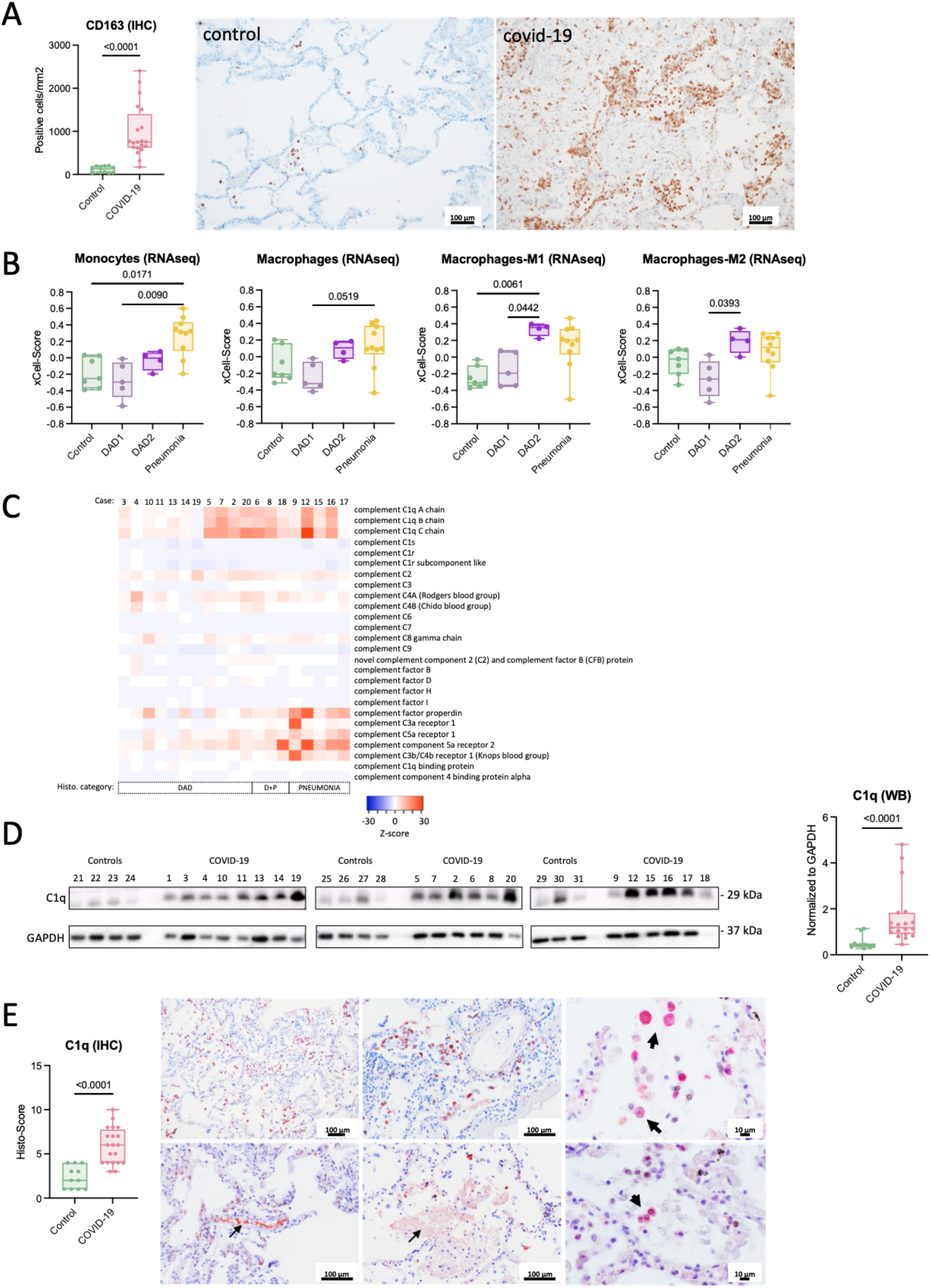
Macrophage and complement C1q induction in covid-19 lungs. (A) Immunohistochemical counting of CD163 positive macrophages shows induction in covid-19 compared to controls (Mann-Whitney test). (B) Both M1 and M2 macrophages are specifically increased in “DAD2” compared to “DAD1” (grouping according to xCell analysis; Kruskal-Wallis test). (C) Heatmap of complement genes specifies C1q induction in a subgroup of DAD cases and in pneumonia. (D) C1q protein (29 kDa) is significantly increased in covid-19 lung tissue compared to controls (reference human GAPDH; Mann Whitney test). (E) Significant Induction of C1q detected by immunohistochemistry (Mann Whitney test) and different staining patterns in covid-19 lungs; top left & middle: C1q staining of alveolar cells; top right: double immunohistochemistry staining (red: C1q, nuclear black: TTF-1) shows C1q staining of alveolar macrophages; bottom left: intravascular C1q staining; bottom middle: free C1q specific staining of proteinaceous fluid in the alveolar space; bottom right: double immunohistochemistry staining (red: C1q, nuclear black: TTF-1) shows C1q staining of pneumocytes (TTF-1 positive).

Among the most discriminative genes between DAD subtypes we found complement factor *C1q* dominantly induced in “DAD2” (Figure 5C). Complement activation is implicated in DAD pathogenesis and linked to severe covid-19 [27-29]. Other complement factors showed no discriminative expression pattern between pathological subgroups in our cohort, except certain complement receptors and properdin mainly induced in pneumonia cases (Figure 6C). Western blots generated from protein extracts of lungs confirmed significantly increased C1q (Figure 6D). A major source of C1q are macrophages corroborated also by a recent single-cell transcriptomic analysis of covid-19 lungs (Figure S11) [30, 31] suggesting a strong connection between macrophages and complement C1q in covid-19 [32]. Immunohistochemical analysis of lung tissues with a C1q antibody showed staining of the vasculature, the interstitial and alveolar space but also of alveolar cells including macrophages and pneumocytes, indicating a multifaceted deposition of C1q in the context of covid-19 in our series (Figure 6E).

C1q is the initiating component of the classical complement cascade but exhibits also immune regulatory functions. It induces the development of pro-resolving M2 type macrophages and is involved in the clearance of apoptotic and necrotic cells, which are highly increased in covid-19 lungs [33-35]. In this process C1q binds to cellular break-down products and is subsequently recognized by phagocyte receptors like the leukocyte-associated immunoglobulin-like receptor 1 (LAIR-1; syn.: CD305) conferring uptake and triggering a tolerogenic state in the phagocyte [36, 37]. As shown by a recent single-cell transcriptomics analysis of covid-19 lungs, *LAIR-1* is mainly present in macrophages (Figure S12) [17]. LAIR-1 together with LILRB4 (leukocyte immunoglobulin-like receptor subfamily B member 4; syn.: ILT3) belong to immunoglobulin-like receptors recognizing collagen domains such as present in C1q, thereby inhibiting immune activation [38, 39]. RNAseq confirmed significant induction of *LAIR-1* and *LILRB4* dominantly in “DAD2” followed by “pneumonia” (Figure 7A). Expressions of all 3 *C1q* polypeptide chains (*A, B & C*) significantly correlated with *LAIR-1* and *LILRB4* expression but not with induced collagens (Figures 7B and S13). This might suggest a functional link between C1q and the immune inhibitory receptors LAIR-1 and LILRB4.

**Figure 7.**
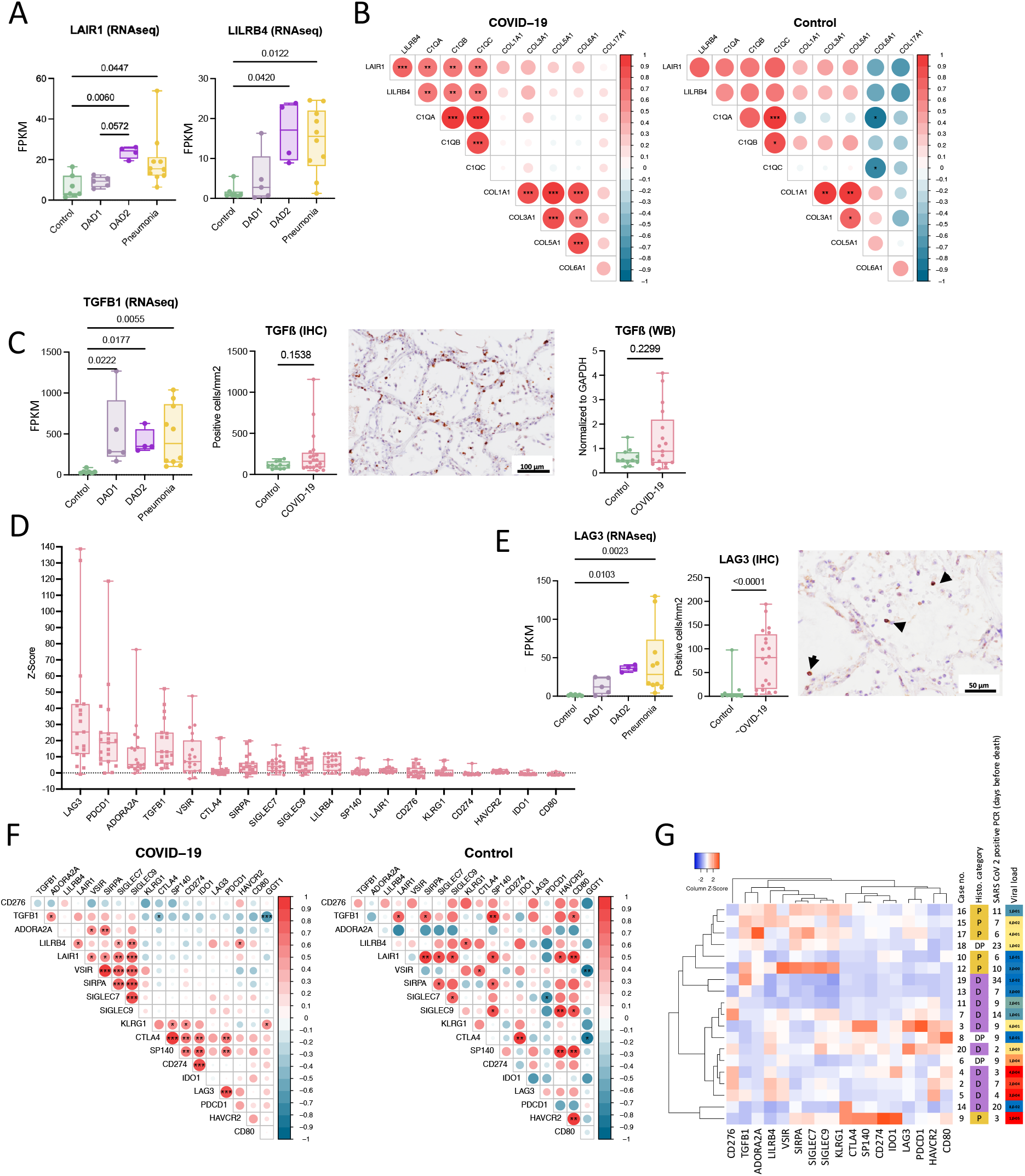
Signatures of immune-impairment in covid-19 lungs. (A) The tolerogenic leukocyte receptors *LAIR-1* and *LILRB4* are mainly induced in “DAD2” and “pneumonia” (Kruskal-Wallis test). (B) Spearman correlation of RNA expression of *LAIR-1* and *LILRB4* with *C1q* chains and induced collagen types (Spearman R; p*<0.05 to p***<0.001). (C) Significant induction of *TGF ≤1* transcription (Kruskal-Wallis test). Protein measurement by immunohistochemistry and Western blotting does not show a significant difference of covid-19 lungs to controls (Mann-Whitney test). (D) Strong induction of immune checkpoint inhibitors in covid-19 (order according to z-score). (E) *LAG3* transcriptional induction (Kruskal-Wallis test) and increased lymphocyte staining with immunohistochemistry in covid-19 lung tissue (Mann-Whitney test). (F) Simultaneous transcriptional induction of immune checkpoint inhibitors in covid-19 lungs compared to controls (Pearson correlation; p*<0.05, p***<0.001). (E) Hierarchical clusteringof immune checkpoint inhibitors of covid-19 cases shows a separate grouping of samples with high viral loads (transcript abundance) versus samples with the histological pneumonia category (clustering: average linkage; distance measure: Pearson).

Since the development of secondary infections is likely driven by local immune-impairment we screened for other anti-inflammatory markers. TGF-β1 is a key factor in the development and healing response of ALI and also implicated in covid-19 lung pathology [40, 41]. *TGF-β1* transcription was significantly increased in covid-19, showing a huge variability, however, TGF-β1 protein measured by immunohistochemistry and western blotting showed no significant induction compared to controls (Figures 7C and S14). Since several immune and non-immune cell types are able to produce TGF-β1, the observed variability in expression might reflect the temporal heterogeneity of lung pathologies in our cohort. Induction of certain inhibitory immune-checkpoints is reported in covid-19 [42-46]. We confirmed transcriptional induction of several inhibitory immune-checkpoints in covid-19 lungs (*LAG3, PDCD1, ADORA2A, VSIR, CTLA4, SIRPA, LAIR1, SIGLEC9, LILRB4, SIGLEC7, HAVCR2*), whereas some showed no induction (*CD276, SP140, IDO1, KLRG1, CD274*) or were even reduced (*GGT1, CD80*) compared to controls (Figures 7D and S15). Noteworthy, the top induced inhibitory immune-checkpoint was found to be *LAG3* (lymphocyte-activation gene 3; syn.: CD223), dominantly induced in “DAD2” and “pneumonia” based on RNAseq, which was also confirmed by immunohistochemistry wherein mainly lymphocytes showed strong staining signals (Figures 7E and S16). LAG3 was recently described as a major increased factor in a plasma proteomic study of severe covid-19 [47]. During immune exhaustion multiple inhibitory receptors act often in synergy amplifying immune impairment, like LAG3 and PD-1 co-induced during chronic viral infections [48]. We confirmed synergistic induction of several inhibitory immune-checkpoints in covid-19 lungs, which showed a different costimulatory pattern compared to controls (Figure 7F). Interestingly, co-expression patterns discriminated cases with high viral loads (*KLRG1, CTLA4, SP140, CD274, IDO1, LAG3, PDCD1, HAVCR2, CD80*) from samples with pneumonia (*CD276, TGFB1, ADORA2A, LILRB4, LAIR1, VSIR, SIRPA, SIGLEC7, SIGLEC9*) suggesting a divergent pattern of induction of inhibitory immune-checkpoints during the course of covid-19 lung pathology (Figure 7G). In summary, these data highlight that multiple pillars of immune impairment act in severe covid-19, leading to a reduced antimicrobial defense in lungs driving the development of secondary infections. The molecular dissection of cell types and immune inhibitory signals might enable the development of specific measures counteracting this potentially lethal complication.

## Discussion

We performed a systematic autopsy study of 20 consecutive covid-19 cases and 14 controls to gain unbiased information about lethal disease courses from the early pandemic. Integration of autopsy, cultivation and deep sequencing provided important clues about host and microbial factors involved in the development of secondary infections as a major sequel of lethal covid-19. Thus, our study might serve as a blue-print for a “holistic” autopsy approach tempting to gain relevant pathophysiological insights from a newly emerging disease. Viral genotyping facilitated directly from autopsy material provided epidemiologic clues about transmission and captured already early events of viral genetic adaptation. Noteworthy, a significant proportion of corpses yielded cultiviable SARS-CoV-2 indicating that autopsy might facilitate virus spread and that special safety requirements should be applied during post-mortem examinations of covid-19 patients [49]. Our investigation showed that covid-19 lung pathology is multifaceted and that a major discriminator of lethal courses is DAD and the presence of secondary infections. This was evident by histology but also mirrored by the deep transcriptomic analysis and microbiology. Secondary infections are reported to develop in up to 42% of patients with covid-19 [7]. Notably, DAD caused by the virus itself and secondary infections are chronologically divergent and provoke overtly different host reactions. Noteworthy, SARS-CoV-2 infection alone might not trigger prominent neutrophil recruitment to the lung at all and neutrophil signatures found in recent covid-19 studies might likely already indicate secondary infections [15, 18, 31, 50, 51]. Thus, it is important to seek for a proper pre-classification of tissue samples based on histology to omit wrong conclusions in omics-based analyses.

The resident lung microbiome is a relevant factor in the pathogenesis of lung infections and reported to be altered in sepsis and DAD [9, 10]. Secondary lung infections are also a complication in influenza, SARS and MERS, wherein bacteria like *S. aureus* or *Klebsiella* spp. and fungi like *Candida* or *Aspergillus* spp. are emerging [13, 14]. Such microbial agents were also prevalent in our cohort. Curiously, the mechanistic understanding why secondary infections develop on top of viral infections is still limited. We could not identify any associated clinical parameter clearly correlated with secondary infections but showed that lung immunity is impaired in covid-19, which might drive these infections. This finding was also underscored by the presence of polymicrobial infections and EBV indicative for a general decreased immunity [12]. Immune exhaustion seems to follow the systemic immune hyperactivation in severe covid-19 and myeloid cells, which are important for the recognition of virus infected cells seem to be key for initiating the proinflammatory response [52, 53]. Recent single-cell transcriptomic studies of covid-19 patients identified myeloid cells as a major induced cell type in BALF specimens with high proportions of proinflammatory macrophages [15, 50]. Generally, M1-type macrophages dominate early DAD, whereas later DAD stages show increased M2-types involved in tissue repair with immunosuppressive features [23]. Thus, later (organizing) phases of DAD might be specifically prone to acquire secondary infections.

Respiratory failure in covid-19 is linked to strong complement activation [27, 28, 54]. Extensive deposition of complement factors, including C1q, in vessels and epithelial cells of lungs and skin was reported in covid-19 [55, 56]. Notably, the SARS-CoV-2 spike-protein might directly activate complement via the alternative pathway [57]. Complement in covid-19 is currently discussed mainly in the context of endothelial injury and fibrin-clot formation [29]. Our study suggests another pathophysiological role, wherein C1q and macrophages might perpetuate immune impairment. Immune complexes formed by viral antigens and antibodies can activate factor C1 as shown in SARS-CoV infection [58]. C1q is involved in the clearance of apoptotic and necrotic cells by phagocytes, a process termed efferocytosis [59]. Apoptosis and necrosis are prominent in covid-19 lungs [33, 34, 47]. During efferocytosis suppression of overwhelming inflammation is important and phagocytes involved in this process are producing anti-inflammatory cytokines. Therefore, C1q binds to molecules released from apoptotic and necrotic cells (e.g. phosphatidylserine, nucleic acids, etc.) and these complexes are recognized by receptors present on phagocytes, like LAIR-1, conferring uptake and inducing a tolerogenic state [30, 35-37]. Noteworthy, binding of C1q to LAIR-1 on plasmacytoid DCs restricts the production of type I interferons impairing antiviral defense, which also occurs in covid-19 [36, 39]. The “DAD2” subtype in our study shows increased macrophages, C1q and LAIR-1 and might therefore represent cases with a lowered immune tone prone for the development of secondary infections. Overall the progression of early (exudative) DAD into late (fibrotic) DAD indicates healing of ALI characterized by significant connective tissue remodeling and a reduced inflammatory tone [60, 61]. Immune suppressive factors such as TGF-β1 are known to be involved in this process and also LAIR-1 and LILRB4 recognizing collagens or collagen-like proteins might act anti-inflammatory during this disease phase [39, 62-65]. Moreover, the synergistic induction of several tolerogenic factors including inhibitory immune-checkpoints [42-47, 66] and increased (apoptotic) cell death of immune-cells [67, 68] altogether perpetuate immune failure in covid-19.

The limitations of our descriptive study are that causalities cannot be directly inferred and that the relatively small cohort cannot show the entire picture of severe covid-19 and associated secondary infections. Varying clinical courses and different comorbidities might also have influenced our findings. In addition, treatment of covid-19 has changed since the early pandemic, thus, current severe courses and developing sequels might also have changed. We also cannot be sure whether the two described forms of DAD might represent just a spectrum of pathophysiological states or are specific pathotypes. Moreover, post-mortem effects like RNA degradation might have introduced additional noise in our investigation. Nevertheless, we found autopsy complemented with microbiology and molecular measures as a powerful tool to gain relevant clues about covid-19 pathophysiology. Importantly, there exists an obvious knowledge gap in the understanding of the molecular mechanisms driving the development of secondary infections on top of in viral lung diseases. This should initiate further studies to understand the molecular pathways in more detail and to unravel chronological phases of immuno-suppression which could also lead to development of rational therapies counteracting this sequel not only in covid-19. For these investigations, autopsy specimens and associated molecular data might serve as a valuable resource.

## Material and Methods

### Autopsy procedure & specimen collection

Autopsies were performed according to CDC guidelines (https://www.cdc.gov/coronavirus/2019-ncov/hcp/guidance-postmortem-specimens.html) and the epidemic response plan of the county of Styria in a BSL-3 facility that has been specifically designed for post-mortem examinations and sample collection [49]. Full autopsies were performed and swabs (eSwab, Copan), tissue and body fluid samples were taken. Tissues were immediately fixed in 10% buffered formalin (for histology) or 2,5% buffered (sodium cacodylate; pH 6.5; Sigma) glutaraldehyde (for electron microscopy), snap frozen in liquid nitrogen or preserved in RNAlater (ThermoFisher) and stored at -80°C until further processing.

### Histopathology and immunohistochemistry

Formalin-fixed paraffin-embedded (FFPE) tissue specimens were processed and stained according to standard procedures. Stains consisted of hematoxylin & eosin (H&E), periodic acid–Schiff (PAS), chromotrope aniline blue (CAB), sirius red, Gomori, Prussian blue, Giemsa and toluidine blue. Chronic renal changes involving the individual renal compartments were scored according to Sethi et al. [69]. Liver fibrosis was scored according to Ishak et al [70]. The following antibodies were used: Anti-SARS-CoV-2 nucleoprotein (NP) antibody (clone ID: 019, dilution 1:100, rabbit IgG; Sino Biological, Beijing; detection-system: Dako REAL TM EnVision TM HRP rabbit/mouse Dako K5007); CD68 (Ventana anti-CD68 (KP-1) monoclonal mouse 790-2931; detection-system: Ventana Ultra View DAB); TTF1 (Cell marque 343M-96 Clone 8G7G3/1 monoclonal mouse 1:200; detection-system: Dako K5007); TGFß1 (Santacruz polyclonal rabbit AB; clone SC-146 1:50; detection-system: Ventana Ultra View DAB); LAG3 (Abcam polyclonal rabbit; clone ab180187 1:5000; detection-system: Dako K5007); C1q (Dako polyclonal rabbit, clone A0136 1:5000; detection-system: Dako K5007); CD163 (Ventana monoclonal mouse, clone MRQ-26 1:50; detection-system: Ventana Ultra View DAB). RNA in-situ hybridization for EBV was performed with the Inform EBER Epstein Barr Virus early RNA kit (Ventana 800-2824) and the detection-system ISH invers blue (Ventana).

### Scoring of histological lung features

From each case multiple specimens from each lobe were taken to account for variations in disease representation (at least 2 specimens per lobe corresponding to 10 specimens per case at least, mean: 13, range: 10-21) and assessed for histopathological features. Features were grouped according to (a) early (exudative) and (b) late (fibrotic) DAD as well as (c) general cellular and tissue reactions in pulmonary pathology and DAD development [21] and consisted of: Interstitial edema/thickening of the alveolo-capillary membranes (non-fibrotic); alveolar edema; hyaline membranes; intra-alveolar (loose) fibrin; scaled of pneumocytes; intra-alveolar proliferation of pneumocytes; alveolar septal fibrosis (early stage organizing DAD); alveolar septal fibrosis (end stage organizing DAD); bronchialisation; squamous metaplasia; virus induced cellular changes in pneumocytes and/or alveolar macrophages; alveolar hemorrhage; interstitial hemorrhage; interstitial hemosidern (free); alveolar hemosiderin (siderophages); fibrin thrombi (capillaries); fibrin thrombi (larger vessels); vascular remodeling (increased density, vascular fissures); intra-alveolar macrophage accumulation; lymphocytes (within alveolo-capillary membranes); lymphocytes (within interstitial space); intra-alveolar (fibro-) cellular infiltrate; lymphocytes (alveolar); megakaryocytes (capillaries); alveolar neutrophils; bronchial neutrophils; a 4-grade scoring system was used to describe the severity of the different pathological features. Score 0 (feature absent), score 1 (feature present in <=33%), score 2 (feature present in <=66%), score 3 (feature present in > 66%). Scores per slides were summed up and a final score (mean value) was calculated.

### Microbial culture and identification

Native lung tissues were transferred into mixing vessels (ProbeAX; AxonLab) containing 5 ml of physiological saline and were homogenized using a dispersion instrument (ULTRA-TURRAX® Tube Drive; AxonLab). The homogenates were inoculated (0.1 ml aliquots) onto aerobic blood agar, MacConkey agar, chocolate agar, and anaerobic blood agar (BD Diagnostic) and into thioglycollate broth (Oxoid). Plates were incubated at 35°C and 37°C aerobically, in an atmosphere containing 5 % carbon dioxide and anaerobically (Genbox anaer, bioMérieux) for up to 14 days, respectively. Cloudy thioglycollate broths were sub-cultured on plates. Colonies were identified using matrix-assisted-laser-desorption-ionization time-of-flight mass-spectrometry (MALDI-TOF MS; Vitek^®^ MS, bioMérieux or MALDI Biotyper™, Bruker) or 16S rRNA gene sequencing [71].

### Virus isolation

Lung tissues and swabs from lung parenchyma were used for cultivation of SARS-CoV-2 (Table S3). After mechanical disruption samples were frozen (−80°C) and thawed (37°C) twice to increase cell lysis and viral release. 2mL OptiPro SFM medium (Gibco) with 4mM L-Glutamine (Gibco) and 1% penicillin-streptomycin (10,000 U/ml; Gibco) were added to the samples. After centrifugation (10 min., 1500 rcf) the supernatants were filtered through a 0.45µm membrane filter (Millipore) and inoculated on Vero CCL-81 cells with OptiPro SFM medium with 4mM L-Glutamine and 1% penicillin-streptomycin in T25 flasks (ThermoFisher). After 3-4 days incubation at 37°C and 5% CO2, the whole cells were mechanically detached with cell scrapers and passaged including the supernatant on to new Vero CCL-81 cells growing in T75 flasks (ThermoFisher). After 1 week the cells were harvested and supernatants were stored after centrifugation (10min., 1500 rcf) at -80°C. Viral load was determined by qRT-PCR as described below.

### RNA extraction

Samples consisted of swabs (eSWAB, Copan), tissues and body fluids, the latter were collected with sterile syringes. Fresh tissues were sampled directly into Magna Lyser Green Beads tubes (03358941001, Roche) pre-filled with 400µl lysis buffer. Tissues were homogenized with a MagnaLyser (Roche) instrument using 6500 rpm for 30 sec. and 3 repetitions. RNA was extracted from 200µl eSwab solution, 200µl liquid sample or tissue homogenate using the Maxwell 16 LEV simplyRNA Blood Kit (AS1310, Promega) according to the manufacturer’s instructions. RNAs from VeroE6 cell cultures were isolated by using the QIAamp Viral RNA Mini Kit (Qiagen) without addition of carrier RNA and transcribed into cDNA with the High-Capacity cDNA Reverse Transcription Kit with RNase Inhibitor (Applied Biosystems) according to manufacturer’s instructions.

### SARS-CoV-2 quantitative RT-PCR

qRT-PCR was performed using a RdRP gene assay and with a probe specific to SARS-CoV-2 [72]. Briefly, primers, probes and 5µl of RNA were added to 10µl of SuperScript III One-Step RT-PCR System with Platinum Taq High Fidelity DNA Polymerase mastermix (ThermoFisher). PCR was performed on a Quantstudio 7 instrument (ThermoFisher) with the following cycling conditions: 55°C for 15 min, 95°C for 3 min; 45 cycles consisting of 95°C for 15sec and 58°C for 30sec. Amplification data was downloaded and processed using the qpcR package of the R project (https://www.r-project.org/). Amplification efficiency plots were visually inspected and Cp2D (cycle peak of second derivative) values were calculated for samples with valid amplification curves. Plots were generated with R using the reshape, tidyverse and ggplot packages. qRT-PCR of virus cultures employed primer sets recommended by the CDC detecting three different regions of the viral nucleocapsid (2019-nCoV N1-F 5’-GAC CCC AAA ATC AGC GAA AT-3’, 2019-nCoV,_N1-R 5’-TCT GGT TAC TGC CAG TTG AAT CTG-3’; 2019-nCoV_N2-F 5’-TTA CAA ACA TTG GCC GCA AA-3’, 2019-nCoV_N2-R 5’-GCG CGA CAT TCC GAA GAA-3’; 2019-nCoV_N3-F 5’-GGG AGC CTT GAA TAC ACC AAA A-3’, 2019-nCoV_N3-R 5’-TGT AGC ACG ATT GCA GCA TTG-3’) and human RNAse P as a control (2019-nCoV_RP-F 5’-AGA TTT GGA CCT GCGAGC G-3’, 2019-CoV_RP-R 5’-GAG CGG CTG TCT CCA CAA GT-3’) or GAPDH (GAPDH_f 5’-CCTCCACCTTTGACGCT-3’, GAPDH_r 5’-TTGCTGTAGCCAAATTCGTT-3’) as control. PCR was performed with the SYBR Green PCR Mastermix (Applied Biosystems) on a Quantstudio 7 instrument (ThermoFisher) with the following cycling conditions: 25°C for 2 min, 50°C for 15 min, 95°C for 10min, 45 cycles consisting of 95°C for 3sec and 55°C for 30 sec.

### Viral genome sequencing

PCR primers spanning the whole genome of SARS-CoV-2 were designed yielding in about 2kb amplicons (Table S9). 2.5µl of RNA were used in three separate RT-PCR reactions as described above with oligonucleotide primers at 400nM concentration with the following cycling conditions: 55°C for 15 min, 95°C for 3 min; 35 cycles consisting of 95°C for 15 sec and 57°C for 3 min; final extension at 72°C for 10 min. PCR products were combined and purified by incubation with 1.8X Ampure XT beads (Beckman Coulter) followed by two washes with 75% ethanol and elution in 30µl water. Amplicons were fragmented to 150-250bp length and Ion Torrent barcode and sequencing adapters were ligated to the fragments using the NEBNext Fast DNA Fragmentation & Library Prep Set for Ion Torren kit (New England Biolabs) according to the manufacturer’s recommendations. Libraries were sequenced on an Ion Torrent S5XL instrument using a 540 Chip Kit and the 200bp sequencing kit (ThermoFisher). Sequences were aligned to the SARS-CoV-2 reference genome (acc. no.: NC_045512.2) using TMAP (v5.10.11) and variants were called with the Torrent Variant Caller (v5.10-12). All called variants were visually inspected and consensus sequences of the viral genomes were generated with bcftools (v1.3.1). Consensus sequences were aligned using clustalw (v2.1), guide trees were visualized in figtree (v1.4.4) and final adjustments were made with Incscape (v0.92). SARS-CoV-2 genomes from our study were uploaded and analyzed with the GISAID SARS-CoV-2 (hCoV-19) database which can be accessed via https://www.gisaid.org/epiflu-applications/next-hcov-19-app/ [73, 74].

### RNA sequencing

Libraries for RNA sequencing (RNAseq) from lung tissues (19 covid-19 cases #2-#20 and 7 control cases #21, #23-#28) were prepped with the KAPA RNA HyperPrep Kit with RiboErase (HMR) for Illumina® platforms (KAPABIOSYSTEMS) according to the manufacturers protocol. Slight modifications from the protocol consisted of a fragmentation step at 65°C for 1min, 12 cycles of PCR, as well as an additional bead cleanup at the end of the prep. Libraries were pooled in two pools of 13 samples each by concentration measured with Qubit (ThermoFisher), followed by a bead-cleanup step and an additional QC with Qubit (ThermoFisher) and BioAnalyzer (Agilent). Sequences were resolved on a NovaSeq 6000 Sequencer (Illumina) with a standard paired-end protocol. RNAseq data were aligned to the human reference genome using STAR [75] (GRCh38 assembly, Ensembl V99 gene models) in 2-pass mode with the following parameters: -sjdbOverhang 100 -outFilterMultimapNmax 20-outFilterMismatchNoverLmax 0.05 -outFilterScoreMin 0 -outFilterScoreMinOverLread 0 -outSJfilterReads Unique -outSJfilterOverhangMin 20 15 15 15 15 -outSJfilterCountUniqueMin 3 3 3 3 -outSJfilterCountTotalMin 3 3 3 3 -outSJfilterDistToOtherSJmin 0 0 0 0 -- outSJfilterIntronMaxVsReadN 100000 -alignIntronMin 20 -alignIntronMax 100000 -- alignMatesGapMax 100000 -alignSJoverhangMin 12 -alignSJstitchMismatchNmax 5 -1 5 5 - alignSJDBoverhangMin 7 -alignSplicedMateMapLmin 0 -alignSplicedMateMapLminOverLmate 0.5 -limitSjdbInsertNsj 5000000 -clip3pAdapterMMp 0.5 -outSAMmultNmax 1 - outSAMmapqUnique 60 -outFilterType BySJout -outSAMunmapped Within -outWigType bedGraph -outReadsUnmapped None SortedByCoordinate -outSAMattrIHstart 1 - twopassMode Basic -chimSegmentMin 8 -chimOutType Junctions WithinBAM SoftClip - chimScoreMin 1 -chimScoreDropMax 20 -chimJunctionOverhangMin 8 - chimSegmentReadGapMax 3 -quantMode GeneCounts -outSAMstrandField intronMotif - outFilterIntronStrands None -chimMainSegmentMultNmax 2 -outSAMattributes NH HI AS nM NM MD jM jI XS ch. Alignment to the virus genome reference NC_045512.2 was performed using bowtie2-2.4.1 [76] on all reads that did not map to the human genome. Read counts on plus-/minus-strand were counted using custom python scripts. Exact positioning of the reads on plus-/minus-strand was done splitting the bam files aligned to NC_045512.2 using samtools -f 0×10 and samtools -F 0×10 (v0.1.19-44428cd) and bedtools genomecov -ibam BAM NC_045512.2 -d (bedtools v2.17.0).

### RNA profiling

Gene counts were determined using HTSeq (v0.12.4) and normalized as fragments per kilobase per million (FPKM) after TMM correction. Gene set variation analysis (GSVA) was performed against a set of immune signatures with xCell [19] and means were calculated per cell type using custom R-scripts. Graphs and analyses were generated using R-3.6.0. Differential gene expression was conducted using edgeR (https://doi.org/doi:10.18129/B9.bioc.edgeR) [77]. Differentially expressed genes were selected with FDR<0.05, logCPM>1, and FPKM>1 in at least 5 samples. Clustering of differentially expressed genes was performed using hclust hierachical clustering and subsequent cutting of the gene tree at R function cutree with h=0.25. Gene set enrichment analysis for clusters was done using the online tool (https://www.gsea-msigdb.org/gsea/msigdb/annotate.jsp) [78] for canonical pathways and FDR<0.05.

### Single cell transcriptomic metanalysis

Selected genes from single-cell transcriptomic metadata from Xu et al [31] and Delorey et al [17] were analyzed with the single-cell atlas database SCovid [79].

### Microbiome analysis based on RNAseq

Microbiome analysis was performed with the following steps using all reads from STAR alignment not mapping to the human reference: quality filtering using fastx -q 30 -p 26 -Q33 (v0.0.13, http://hannonlab.cshl.edu/fastx_toolkit/), cleaning of the fasta file using seqclean-x86_64 -N -M -A (https://sourceforge.net/projects/seqclean/), realigning to the human reference using blastn against all databases and removal of all reads with 94% similarity. Remaining reads were annotated using MetaPhlAn2 (v2.6.0) [80] and Pathseq (GATK v4.1.0.0) [81] with default settings.

### Microbiome analysis based on the 16S rRNA gene and internal transcribed spacers (ITS)

Bacterial (16S rRNA gene) and fungal (ITS) microbiome analysis from lung tissue was done from FFPE samples which enabled us to preselect samples based on histology showing unambiguous pathology (i.e. DAD vs. pneumonia). DNA was extracted from FFPE tissues using the Maxwell 16 Tissue DNA Purification Kit (Promega). DNA concentration was measured by Picogreen fluorescence. The variable V1–V2 region of the bacterial 16S rRNA gene was amplified with PCR from 50ng DNA using oligonucleotide primers 16s_515_S3_fwd: 5’-TGCCAGCAGCCGCGGTAA-3’ and 16s_806_S2_rev: 5’-GGACTACCAGGGTATCTAAT-3’. This 16S rDNA region was chosen since it gives robust taxonomic classification and has been shown to be suitable for community clustering. Likewise, fungal ITS sequence was amplified with primers ITS1 5’-TCCGTAGGTGAACCTGCGG-3’ and ITS2 5’-GCTGCGTTCTTCATCGATGC-3’. DNA was amplified with the 16s Complete PCR Mastermix Kit (Molzym). The first PCR reaction product was subjected to a second round of PCR with primers fusing the 16s/ITS primer sequence to the A and P adapters necessary for Ion Torrent sequencing while additionally including a molecular barcode sequence to allow multiplexing of up to 96 samples simultaneously. PCR products were subjected to agarose gel electrophoresis and the band of the expected length (330nt) was excised from the gel and purified using the QiaQick (Qiagen) gel extraction system. DNA concentration of the final PCR product was measured by Picogreen fluorescence. Amplicons from up to 60 samples were pooled equimolarly and sequencing was performed on Ion Torrent XL benchtop sequencer using the Ion 400BP Sequencing chemistry (all reagents from ThermoFisher). Sequences were split by barcode and transferred to the Torrent suite server.

Raw bam-files comprised of single-end reads generated by NGS, were converted from bam files to fastq.gz files by using samtools (http://www.htslib.org/). Quality control and preprocessing of sequences was performed using FastQC (version 0.7), MultiQC (version 1.7) and trimmomatic (version 0.36.5) using following parameters: LEADING:3 TRAILING:3 SLIDINGWINDOW:4:15 MINLEN:200. Sequence processing and microbiome analysis was performed using QIIME2 (version 2020.6) [82]. After quality filtering all samples with less than 9833 reads/samples were excluded from downstream analysis. In concordance with RNA-Seq analysis Covid-1 was excluded for sub-analysis (Cause of Death: Myocardial Infarction), resulting in 18 Covid and 12 control samples for 16S analysis (Average frequency: 28201.7 reads/sample). For ITS analysis only 1 sample showed more than 9833 reads/per sample (Covid-18: 10272 reads). All other samples showed no clear ITS signal with a sequencing depth of maximum 219 reads per sample. Denoising, dereplication and chimera filtering of single-end reads were performed using DADA2 (denoise-pyro) [83]. 16S-based analysis was performed with the latest SILVA 138 taxonomy and the Naive Bayes classifier trained on Silva 138 99% OTUs full-length sequences. For ITS-based analysis a classifier was trained on the UNITE reference database (ver8-99-classifier; 04.02.2020) according to John Quensen (http://john-quensen.com/tutorials/training-the-qiime2-classifier-with-unite-its-reference-sequences/; assessed 20/08/2020). Differences in microbial composition between groups were tested with implemented QIIME2 plugins using PERMANOVA (p<0.05, qiime diversity beta-group-significance: Bray-Curtis, Jaccard, Unweighted UniFrac, Weighted UniFrac) and Kruskal-Wallis (p<0.05, qiime diversity alpha-group-significance: Observed features, Shannon, Evenness, Faith PD). For metagenomic biomarker discovery taxonomic feature-tables were introduced to LEfSe (linear discriminant analysis effect size) method (Galaxy version 1.0; p<0.05, LDA>2, All-against-all) [84]. Plots were generated with R (version 3.6.2)6 in RStudio (1.1.463)7 using following packages: tidyverse (1.3.0)8, qiime2r (0.99.6)9, ggplot2 (3.3.3)10, dplyr (1.0.6)11 and ggpubr (0.4.0.999)12 and GraphPad Prism. The graphical abstract was created with BioRender (www.BioRender.com).

### Protein isolation and western blot

Proteins from lung tissues were extracted with TRIzol® (ThermoFisher) according to the supplier’s protocol. Briefly, tissue homogenates were subjected to phase separation wherein the organic phase containing the protein was further processed. Four volumes of ice-cold acetone were added to the organic phase and the mixture was incubated at -20°C overnight, followed by a centrifugation step (13000 rpm) at 4°C for 15 min. The supernatant was discarded and the pellets were dried at 60°C for 60 min. Subsequently 100 µl RIPA buffer (Sigma) containing protease inhibitors and phosphatase (0.1 mM Pefabloc, 1 mM DTT, 1X cOmplete™ Mini, 1X PhosSTOP™) and 1 % SDS (Roche) were added and the mixture incubated at 65°C for 90 min. Supernatants were transferred to a new Eppendorf tube and 100 µl 8M urea in 0.05M Tris (pH 8,5) and 1% SDS were added and incubated at 55°C for 30 min. Corresponding supernatants and pellets were pooled and transferred to 2 ml MagNA Lyser tubes (Roche) with ceramic beads and homogenized 2 times at 6500 rpm for 20 sec. Samples were incubated on ice for 10 min and subsequently centrifugated with 13000 rpm at 4°C for 15 min. Supernatants were transferred to new Eppendorf tubes. For western blotting proteins were mixed with 4X Laemmli buffer (Bio-Rad) and incubated for 10 min at 95°C and then loaded onto 11% (v/v) SDS-PAGE gels (Amersham) and electrophoresed at 80 mA for 2 h and subsequently transferred onto nitrocellulose membranes (Amersham). Blotting efficiency was determined with Ponceau staining (Ponceau S solution, Sigma). Non-specific binding was blocked with 5% (w/v) non-fat dry milk (Bio-Rad) in TRIS-buffered saline and 0.1% (v/v) Tween 20 (Merck) for 1h. Subsequently, the membranes were incubated with antibodies against C1q (Dako Denmark A/S 1:5000), TGFß1 (Cell Signaling Technology, 1:1000), and GAPDH (Cell Signaling Technology, 1:1000) overnight at 4°C. Thereafter, membranes were washed and incubated with the appropriate HRP-conjugated secondary antibody (Amersham, ECL Anti-Rabbit IgG, 1:5000). Immunolabeling was detected using ECL Select Western Blot Reagent (Amersham) and visualized with the ImageQuant™ LAS 500 (Amersham). GAPDH was used as loading control to determine protein abundance and band density was quantified and compared by using ImageJ.

## Data Availability

All data produced in the present work are contained in the manuscript and supplemental material; nucleic acid data is deposited in ENA under the acc. no. PRJEB45873.

https://www.ebi.ac.uk/ena/browser/home

## Ethics statement

The study was approved by the ethics committee of the Medical University of Graz (EK-number: 32–362 ex 19/20).

## Data availability

The sequencing data has been deposited in ENA under the acc. no. PRJEB45873.

## Acknowledgements

We are grateful to Tanja V. Mascher, Birgit Gangl, Margit Gogg-Kamerer, Iris Kufferath, Sylvia Eidenhammer, Christine Langner, Stella Wolfgruber, Daniela Pabst, Helmut Donnerer and Lajos Redecsi for their technical assistance.

## Funding

Supported from the Medical university of Graz and the Austrian Science fund (FWF, DK-MOLIN W1241). The design of the BSL-3 laboratory used for autopsy of covid-19 cases was supported by the EU-funded program “European Research Infrastructure for Highly Pathogenic Agents” (ERINHA-Advance, Grant agreement 824061).

## Author contributions

Conceptualization and methodology, MZ, GG, KK, PR, KZ. Investigation and formal analysis, MZ, KK, PW, PR, MS, MN, SE, LM-M, GK, ML, AB, EL, AT, EW, SS, M-JP, F-RV, CL, BJ, LO, GG. Resources, KZ, PR, GG, BT, HL. Writing of original draft GG. Writing, review and editing, all authors. Funding acquisition, KZ, GG, HL. Supervision, GG, PR, KZ. All authors have read and agreed to the published version of the manuscript.

## Supplemental Figures

**Figure S1.**
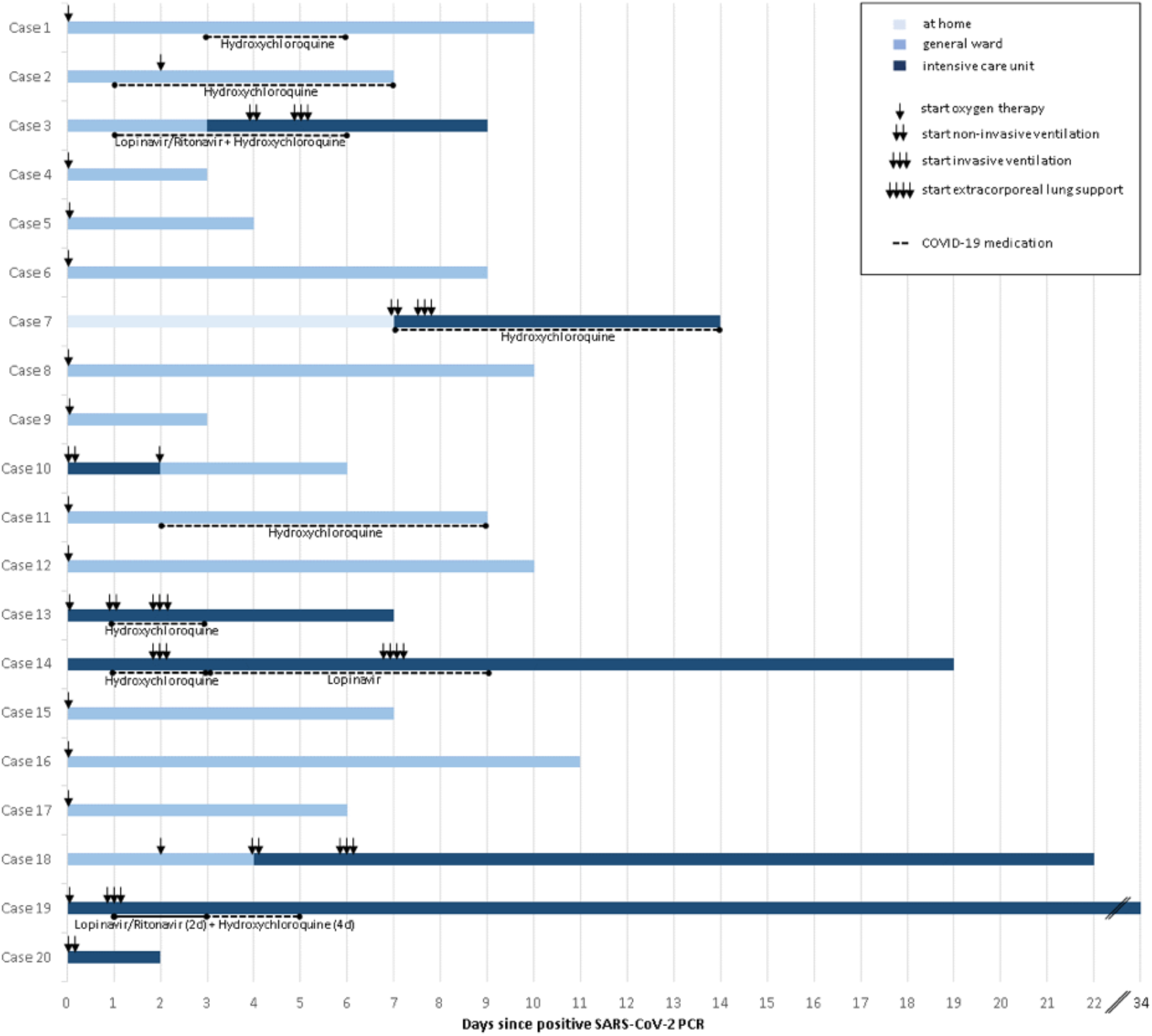
Clinical courses of covid-19 cases. X-axis specifies interval from SARS-CoV-2 positive PCR. Treatment in the general ward and/or intensive care unit is shown. Start of ventilation therapies is indicated by arrows. Covid-19 specific therapies are indicated.

**Figure S2.**
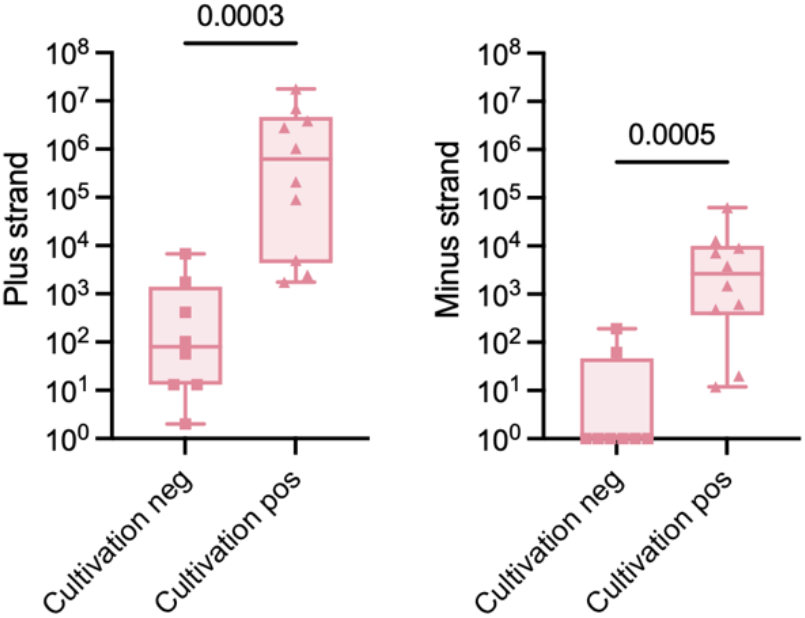
Relation of SARS-CoV-2 cultivability and transcript levels determined by RNAseq (Mann-Whitney test).

**Figure S3.**
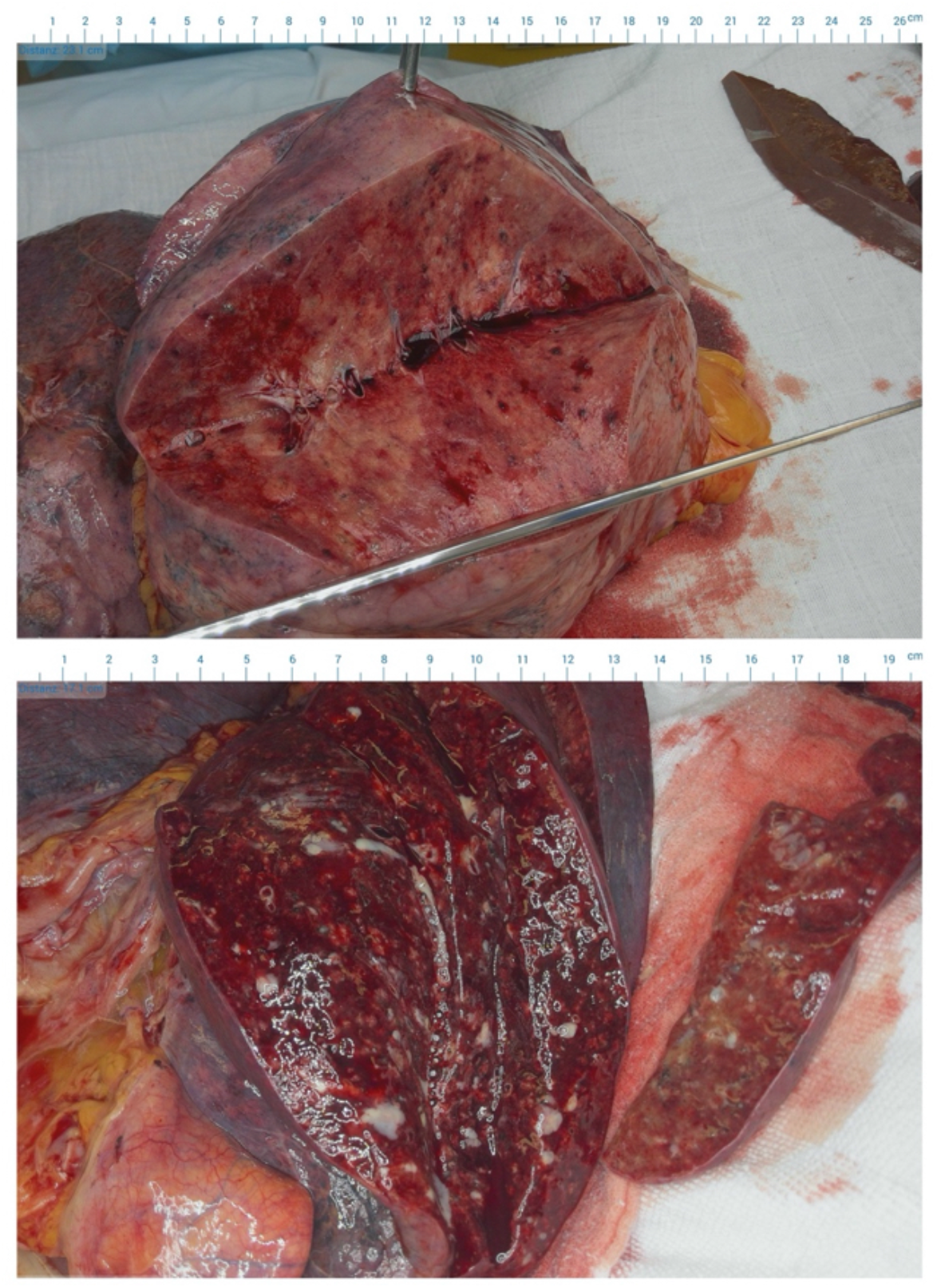
Gross pathology representation of lung. **(Top)** Section through a lobe with DAD. The lung parenchyma is inhomogeneously colored and consolidated. **(Bottom)** A case with bacterial superinfection shows pus in bronchial lumina and on the cut surface of the lung.

**Figure S4.**
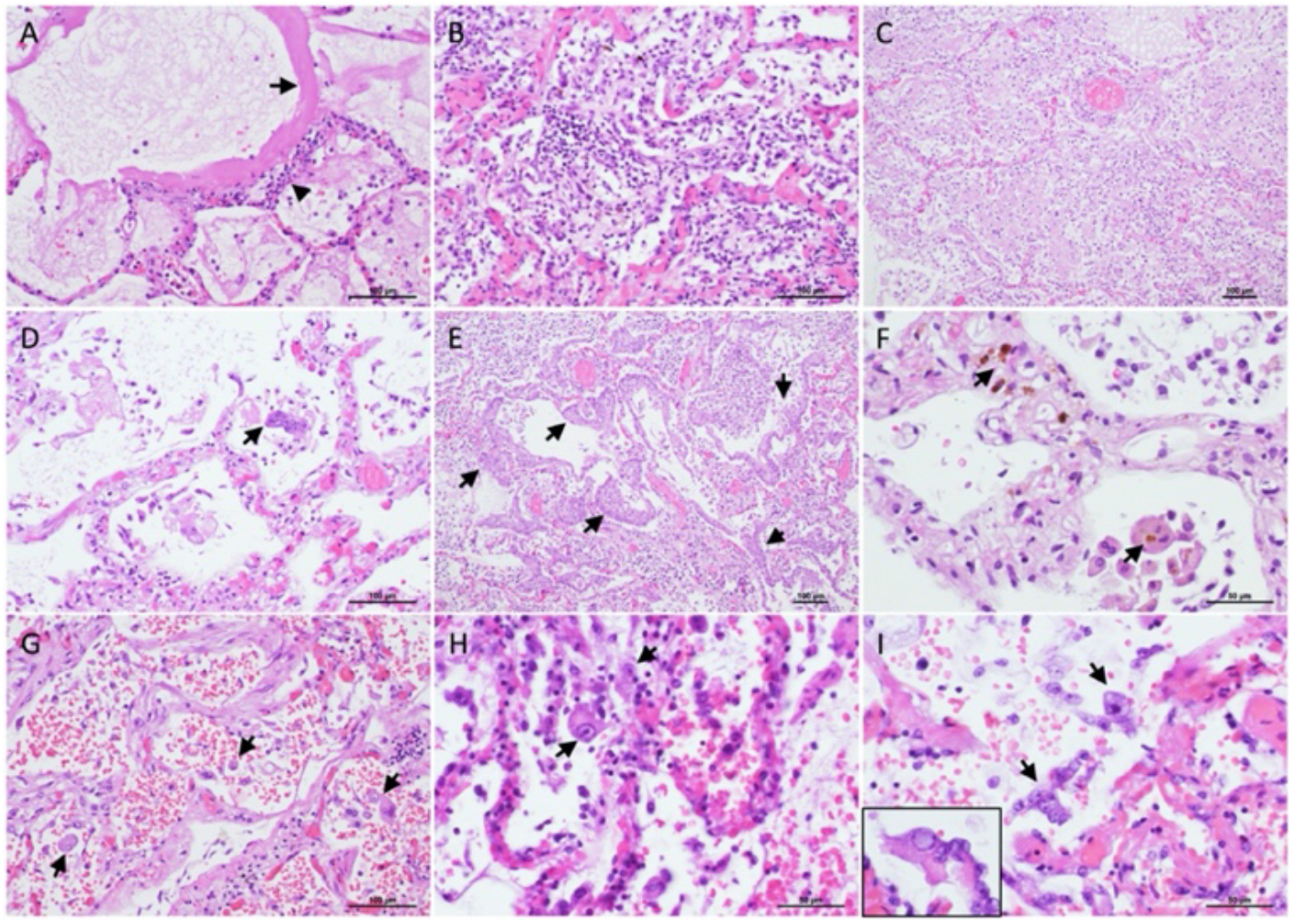
Histological representation of DAD in covid-19. **(A)** Exudative DAD with hyaline membranes (arrows) and lymphocytic interstitial infiltrates in alveolar septa (arrowhead). **(B, C)** Organizing DAD with intra-alveolar fibro-cellular infiltrates. **(D)** Multinucleated syncytial pneumocytes in organizing DAD (arrow). **(E)** Squamous metaplasia in organizing DAD (arrows). **(F)** Hemosiderin in alveolar septa and alveolar macrophages (arrows). **(G-I)** Pneumocyte nuclear atypia with macro-nucleoli (arrows; “owl-eye” in H). Atypical pneumocytes are often scaled off the alveolar membrane. In G also alveolar hemorrhage is evident. Inset in I shows ground-glass nuclear changes in atypical pneumocytes.

**Figure S5.**
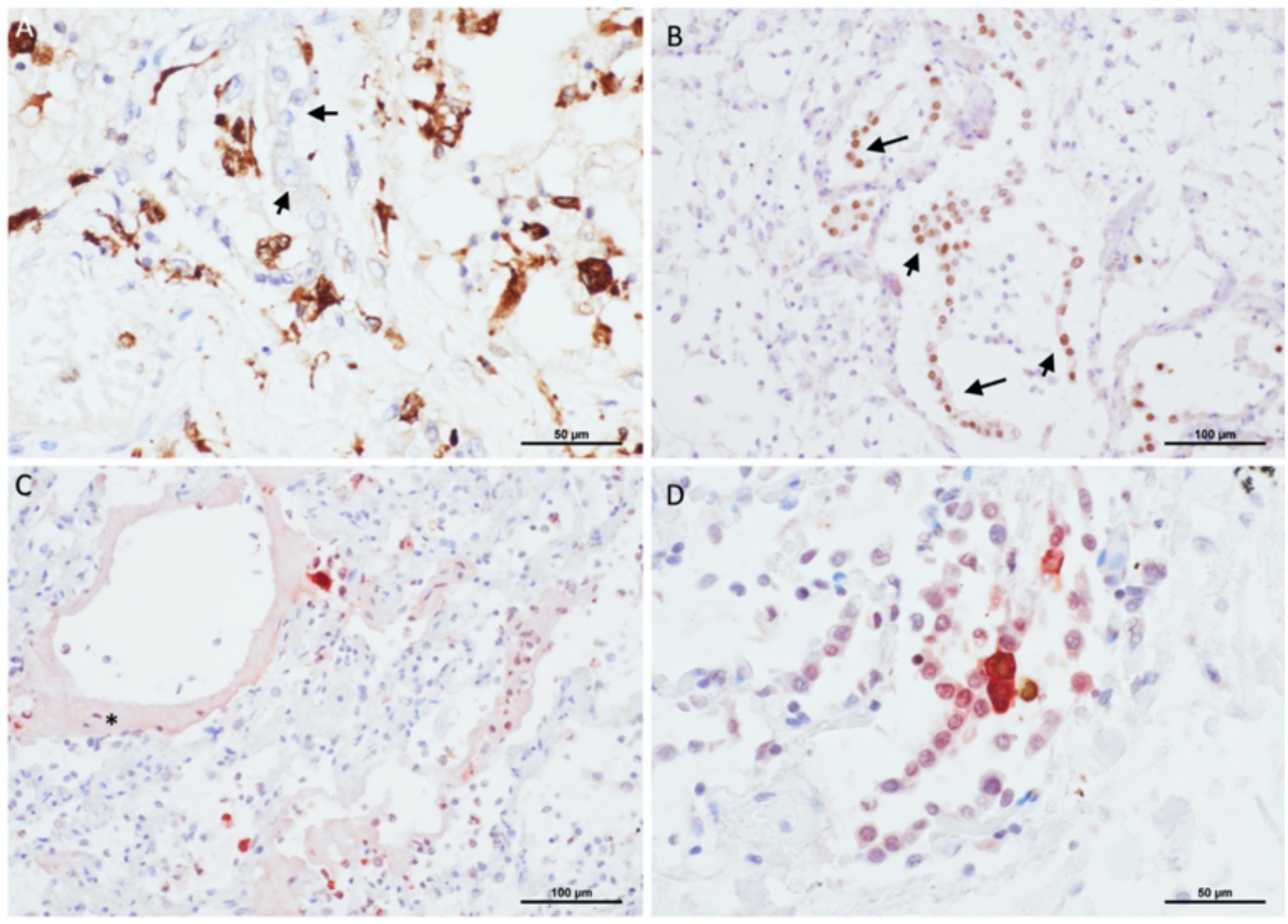
Immunohistochemical analyses of DAD. **(A)** Macrophage specific CD68 staining. Note the non-stained atypical pneumocytes (arrows). **(B)** Pneumocytes are scaled-off from the alveolar membrane (pneumocyte specific nuclear TTF-1 staining, arrows). **(C, D)** SARS-CoV-2 nucleoprotein staining of infected pneumocytes; asterisk (*) marks a hyaline membrane.

**Figure S6.**
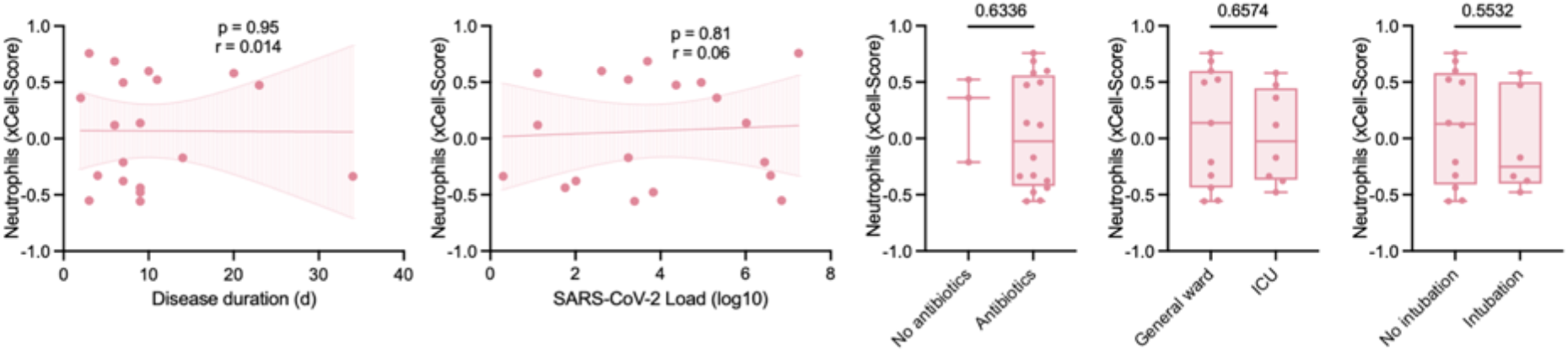
Neutrophils and clinical parameters. Correlation analyses of neutrophil abundance (determined by deconvolution of RNAseq data with xCell) and clinical parameters (Spearman correlation, Mann-Whitney test).

**Figure S7.**
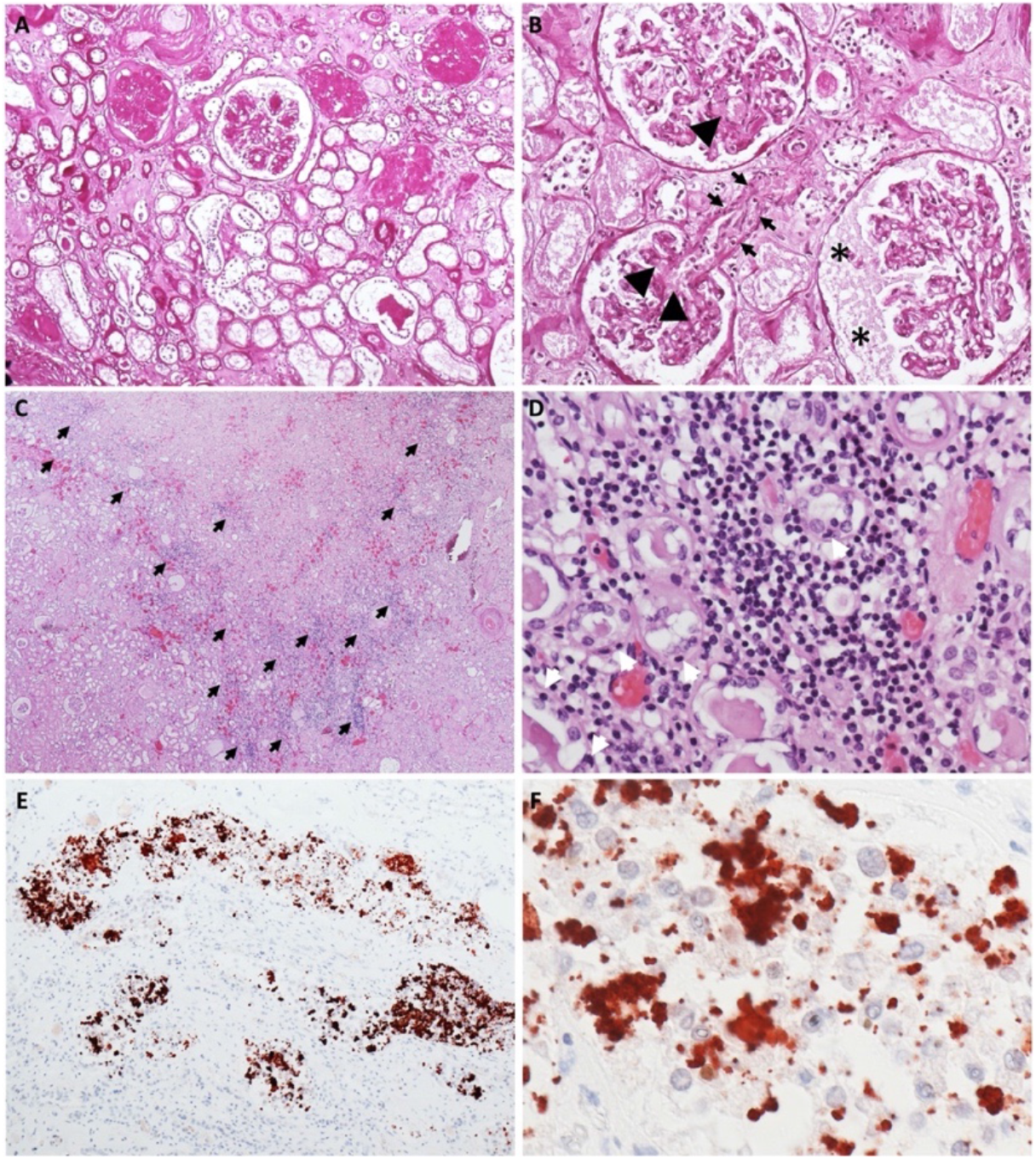
Kidney histopathology. **(A)** PAS-stained kidney specimen with advanced diffuse and nodular diabetic glomerulosclerosis, advanced parenchymal atrophy and severe arteriolohyalinosis (100-fold). **(B)** PAS-stained kidney specimen showing segmental obliteration of glomerular capillary loops by fibrin thrombi (arrowheads), endothelitis of a glomerular arteriole (arrows) and accumulation of plasma in Bowman’s space (asterisks; 200-fold). **(C)** Overview of an H&E-stained kidney specimen with diffuse lymphocytic tubulo-interstitial nephritis (arrows; 20-fold). **(D)** H&E-stained kidney specimen showing severe tubular epithelial changes: loss of brush border, nuclear swelling and cytoplasmic vacuolisation. Note the tubulo-interstitial inflammatory cell infiltrate, mainly consisting of lymphocytes with foci of tubulitis, as characterized by the presence of mononuclear cells on the basolateral aspect of the tubular epithelial cells (400-fold). **(E, F)** Immunohistochemical detection of SARS-CoV-2 nucleoprotein showing granular staining predominantly in the cytoplasm and occasionally in the nuclei of tubular epithelial cells of large distal tubules (100-fold & 600-fold, respectively).

**Figure S8.**
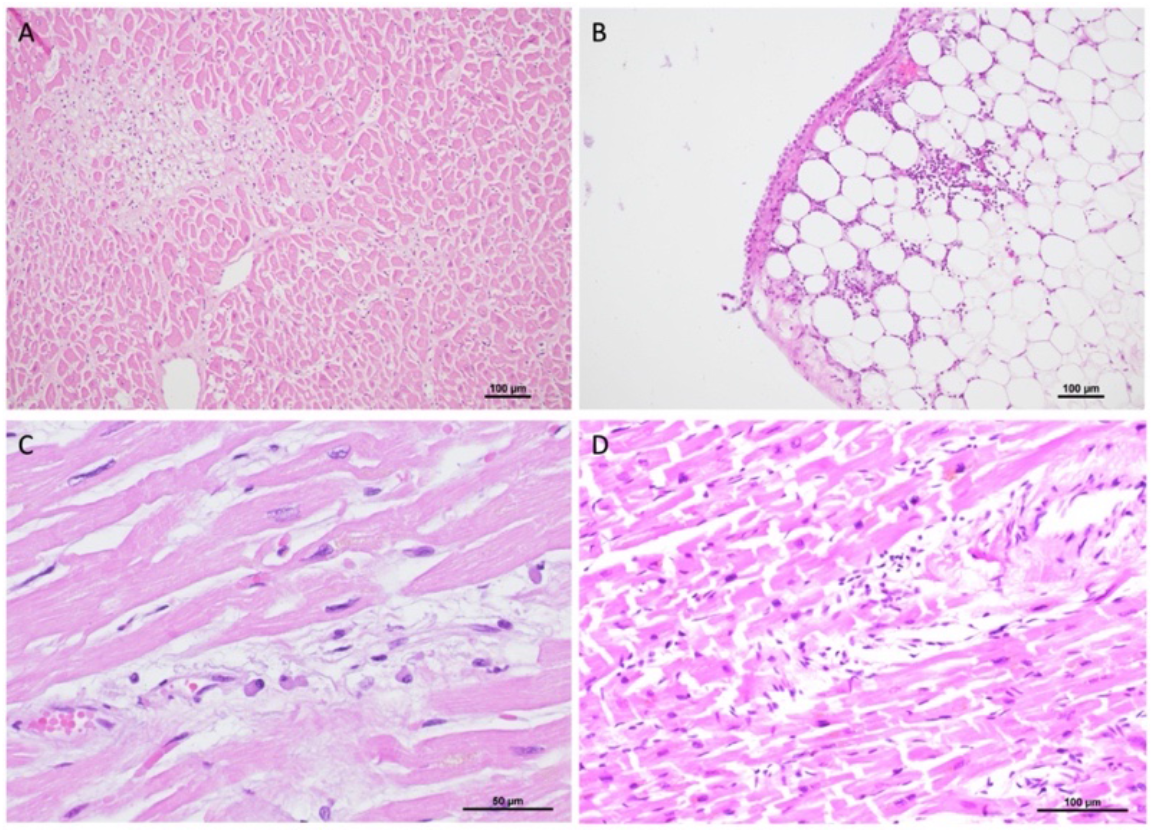
Heart histopathology. **(A)** Fresh small scar in the myocardium. **(B)** Sparse subepicardial lymphocytic infiltrate. **(C, D)** Single plasma cells and lymphocytes in the endomysium around vessels.

**Figure S9.**
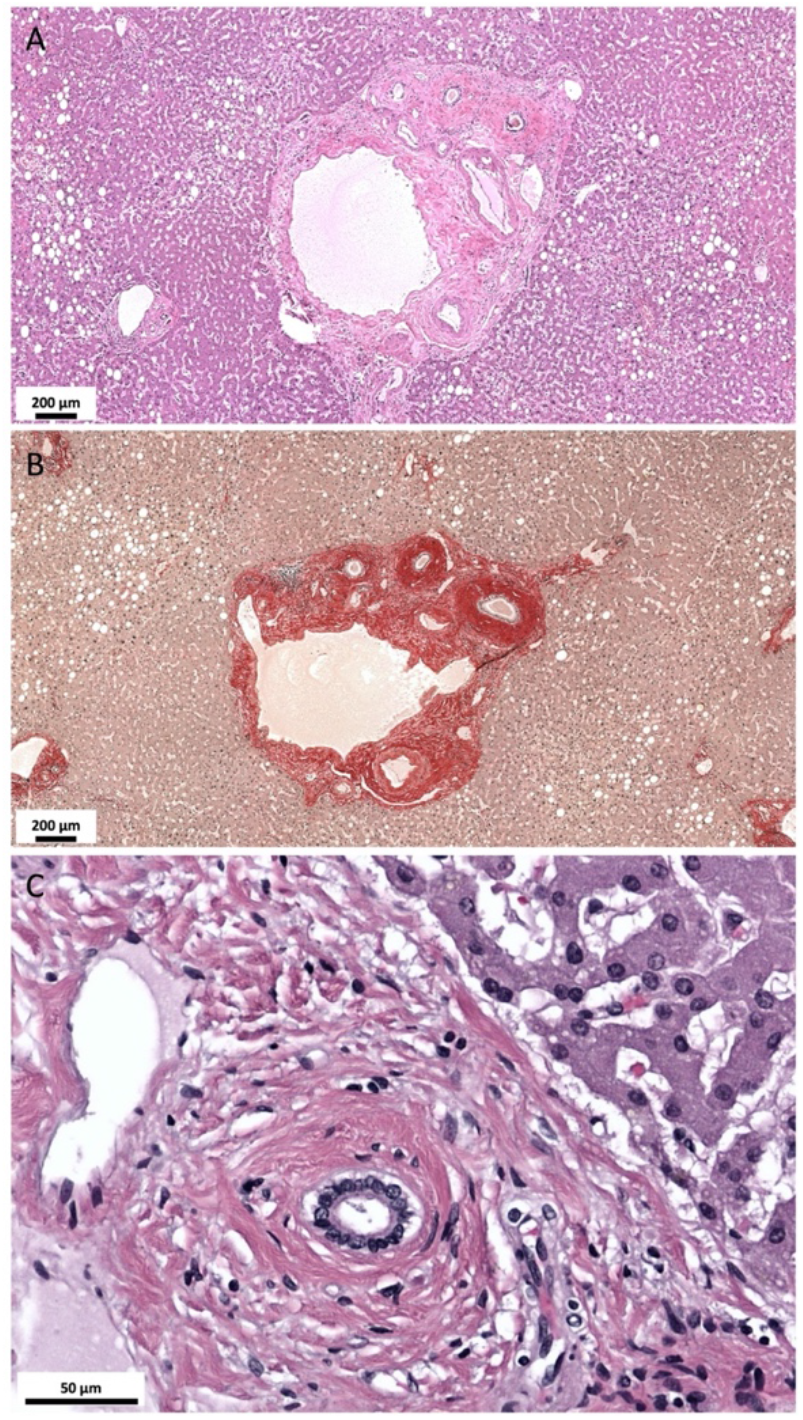
Liver Histopathology. Morphological changes resembling sclerosing cholangitis COVID 19. **(A)** Preserved lobular architecture. Mild macrovesicular steatosis of the lobular parenchyma. Liver septum with mild lymphocytic infiltrates and septal bile duct adjacent to the A. hepatica branch surrounded by thick layer of condensed collagen fibers (50-fold; hematoxylin & eosin). **(B)** Sclerosis is highlighted by the sirius red connective tissue stain (50-fold; sirius red). **(C)** Portal tract with interlobular bile duct and periductal sclerosis.

**Figure S10.**
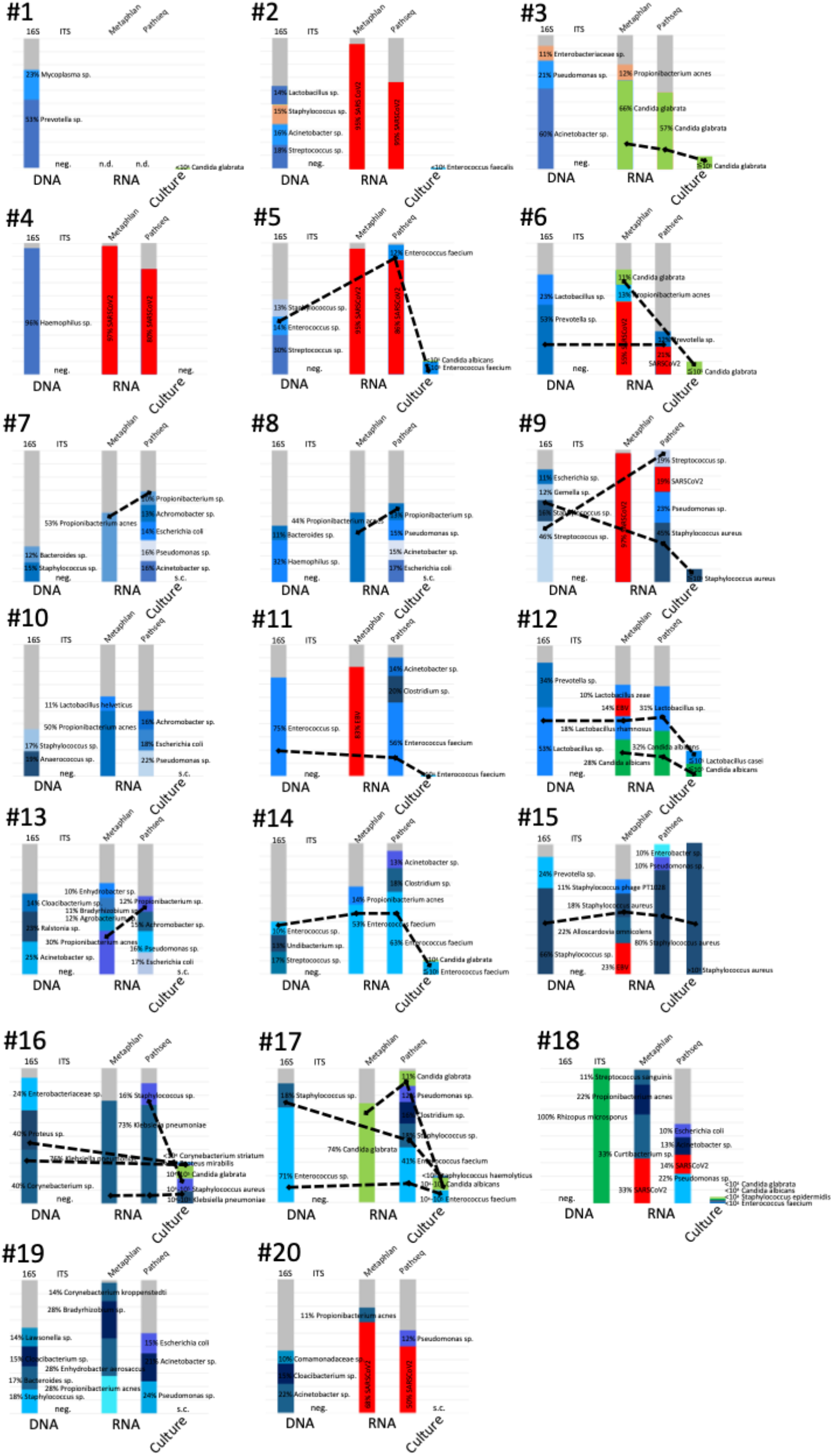
Dominant taxa identified by molecular methods and cultivation in covid-19 lungs. Shown is the relative abundance of 16S rRNA gene and ITS sequencing and microbial RNAseq data annotated with Metaphlan or Pathseq pipelines.

**Figure S11.**
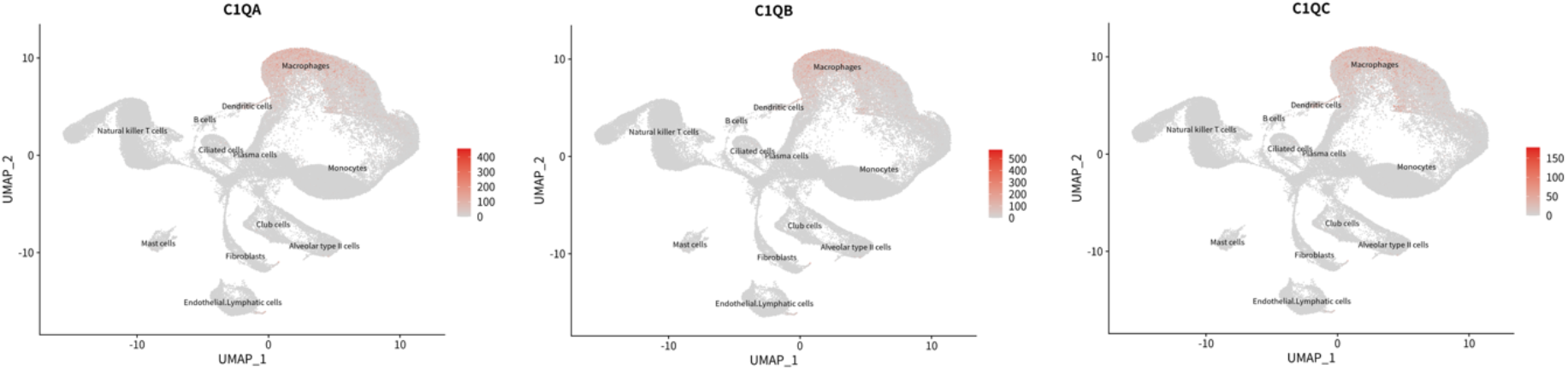
Single cell transcriptomic analysis. *C1q A, B* and *C* chains are mainly expressed by macrophages. Data derived from Xu et al [31] and analyzed with SCovid single-cell atlas database [79].

**Figure S12.**
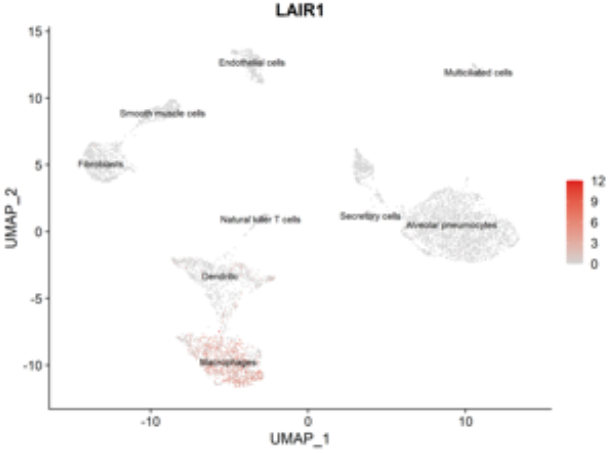
Single cell transcriptomic analysis. *LAIR-1* is mainly expressed by macrophages. Data derived from Delorey et al [17] and analyzed with SCovid single-cell atlas database [79].

**Figure S13.**
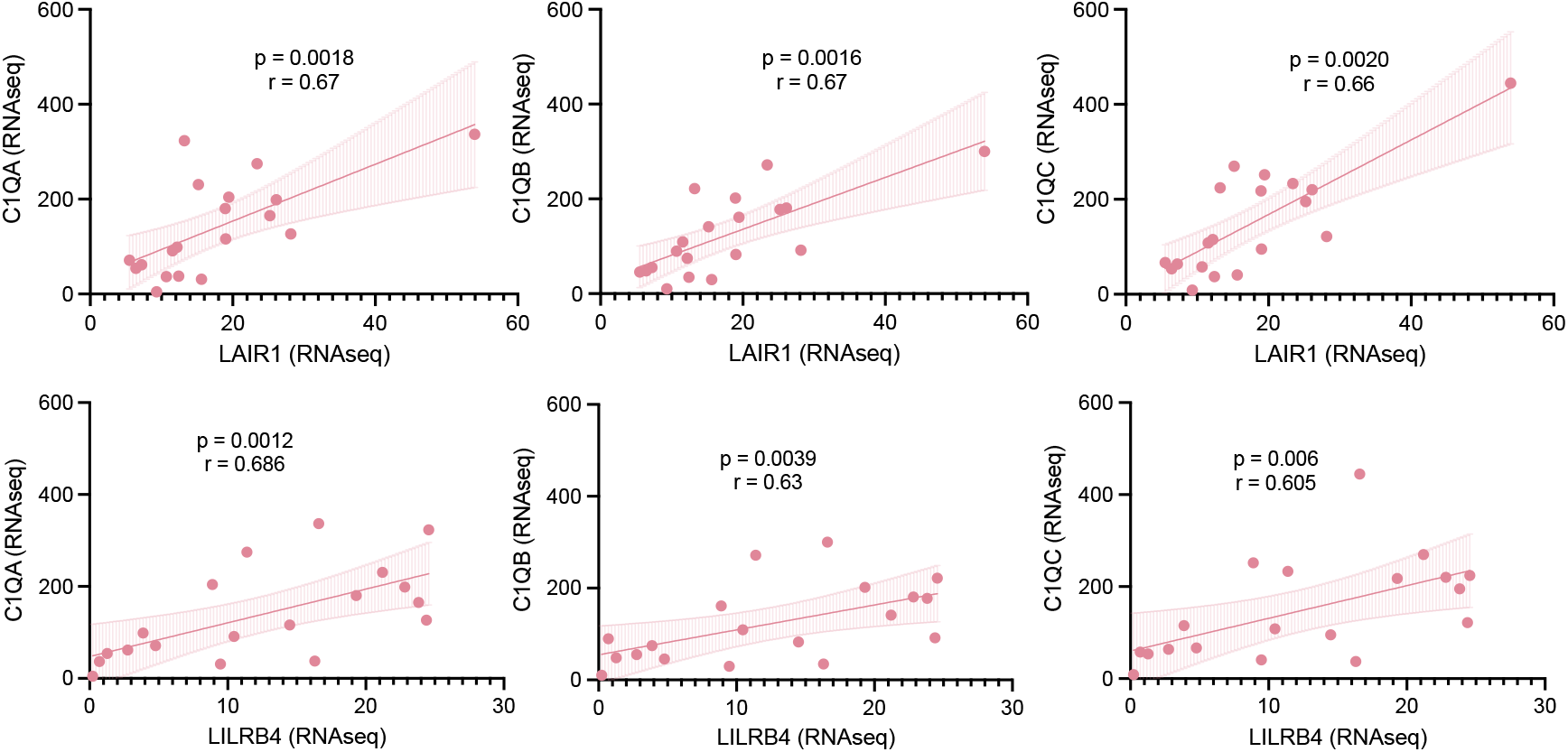
Transcriptomic correlation of *C1q, LAIR-1* and *LILRB4* from RNAseq. Correlation of *C1q* chains with *LAIR-1* (top) and *LILRB4* (bottom; Spearman correlation).

**Figure S14.**
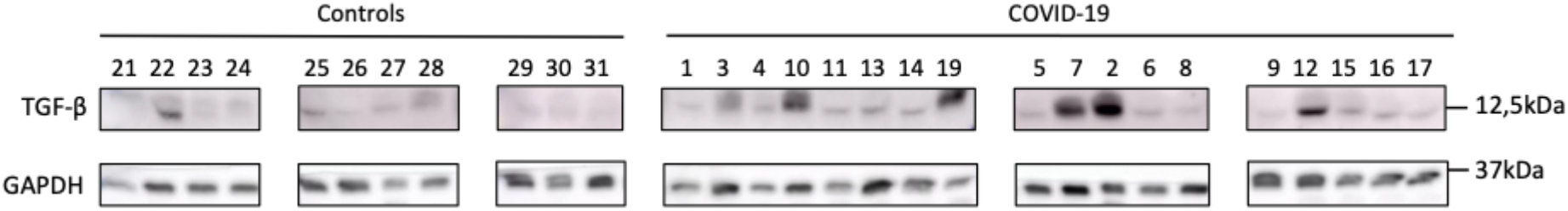
TGF beta 1 (12.5 kDa) western blots in covid-19 lung tissues and controls (reference human GAPDH).

**Figure S15.**
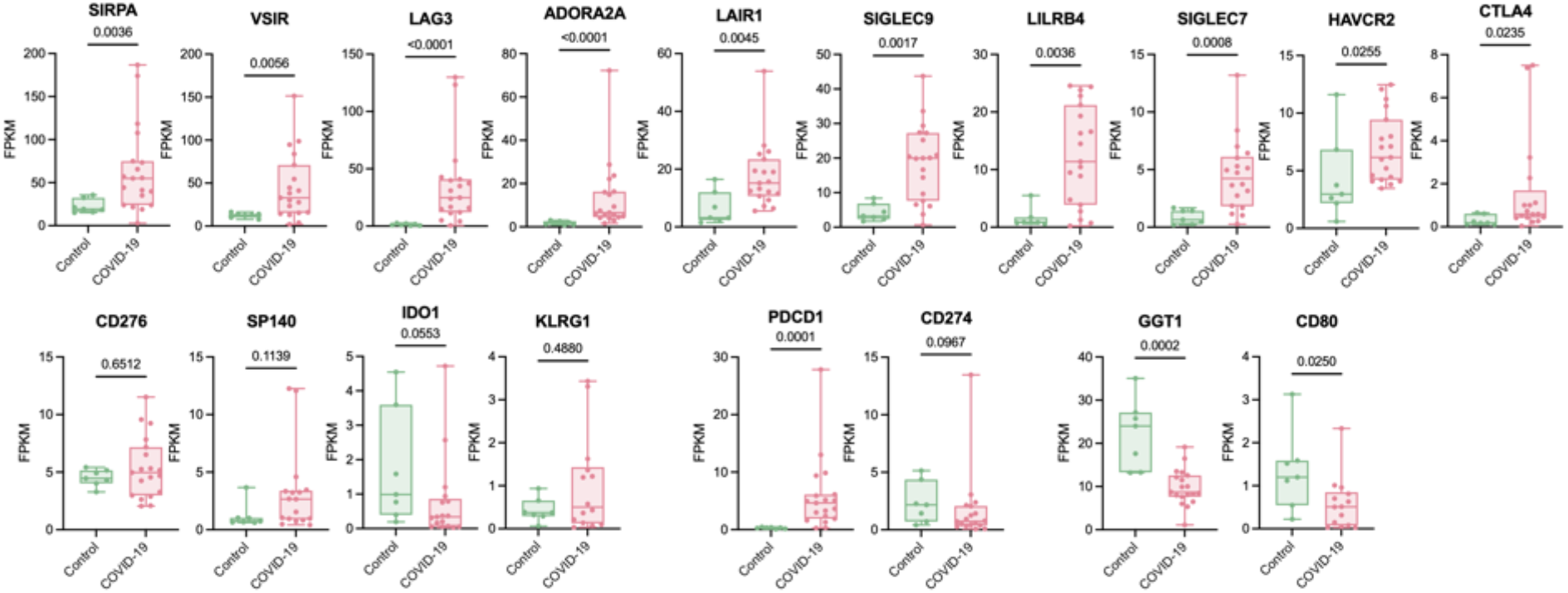
Inhibitory immune checkpoints in covid-19 and controls from RNAseq. Immune checkpoint inhibitor expression in covid-19 compared to controls (Mann-Whitney test).

**Figure S16.**
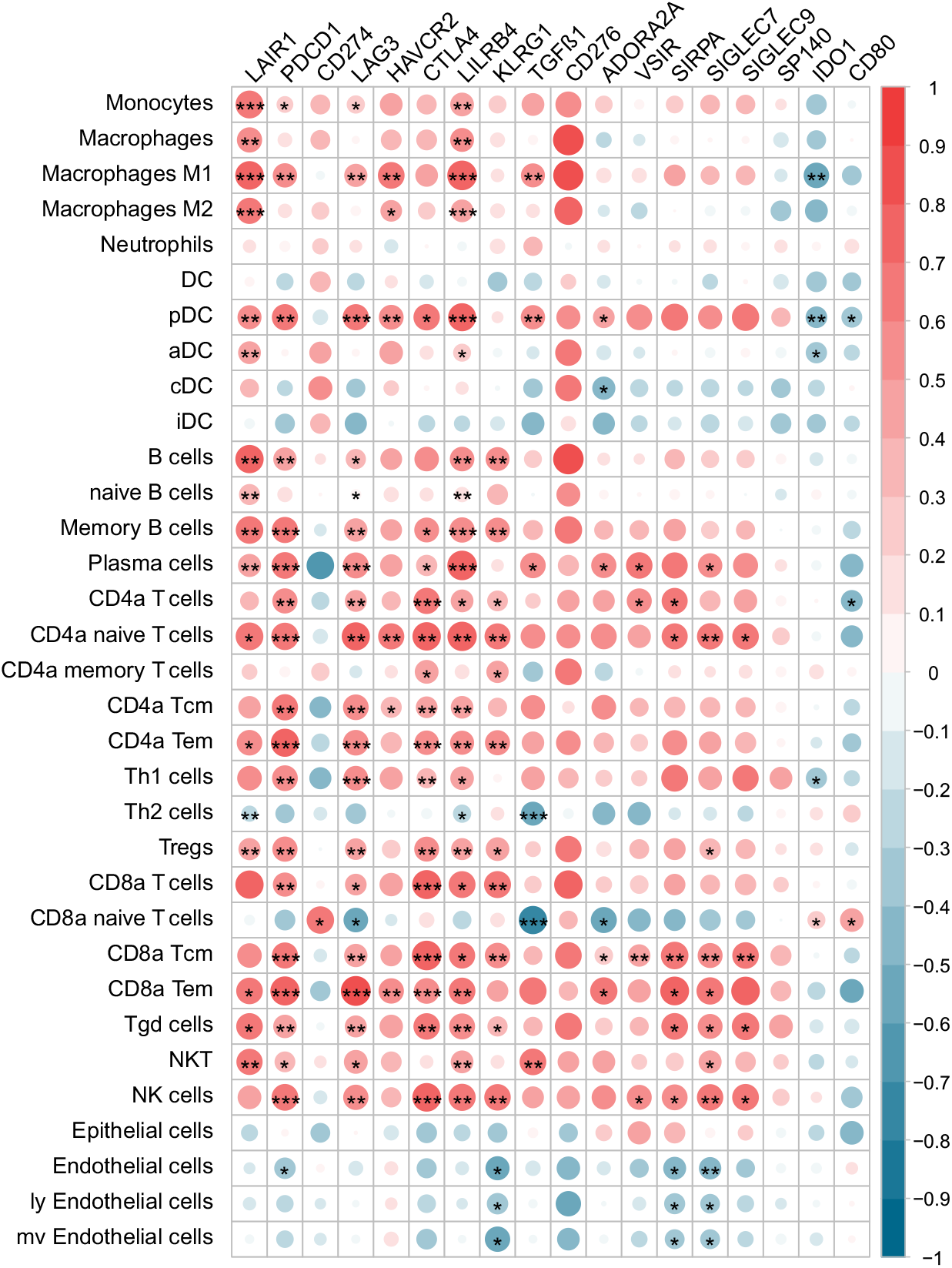
Inhibitory immune checkpoints and cell types from RNAseq data. Correlation analysis of immune-checkpoints and cell types derived from xCell analysis (Spearman correlation; p*<0.05, p***<0.001).

## Notes

### Competing Interest Statement

The authors have declared no competing interest.

### Funding Statement

Support from the Austrian Science fund (FWF, DK-MOLIN W1241). The design of the BSL-3 laboratory used for autopsy of covid-19 cases was supported by the Eu-funded program "European Research Infrastructure for Highly Pathogenic Agents" (ERINHA-Advance, Grant agreement 824061).

### Author Declarations

The study was approved by the ethics committee of the Medical University of Graz (EK number: 32_362 ex 19/20)

